# Immunogenicity and Safety of Gamma, Omicron BA.4/5 and Bivalent SARS-CoV-2 RBD-based Protein Booster Vaccines in Adults Previously Immunized with Different Vaccine Platforms: a Phase II/III, Randomized, Clinical Trial

**DOI:** 10.1101/2024.05.06.24306575

**Authors:** Gonzalo Perez-Marc, Lorena M. Coria, Ana Ceballos, Juan Manuel Rodriguez, Mónica E. Lombardo, Laura Bruno, Federico Páez Córdoba, Clara G. Fascetto Cassero, Melina Salvatori, Mayra Rios Medrano, Fabiana Fulgenzi, María F. Alzogaray, Analía Mykietiuk, Ignacio Leandro Uriarte, Nicolás Itcovici, Tomás Smith Casabella, Gonzalo Corral, Miriam Bruno, Oscar Roldán, Sebastián A. Nuñez, Florencia Cahn, Gustavo A. Yerino, Alejandra Bianchi, Virginia Micaela Braem, Analía Christmann, Santiago Corradetti, Martín Claudio Darraidou, Lucila Di Nunzio, Tatiana Belén Estrada, Rocío López Castelo, Carla Graciela Marchionatti, Lucila Pitocco, Virgina Macarena Trias Uriarte, Cristian Jorge Wood, Romina Zadoff, Florencia Bues, Rosa M. Garrido, Laboratorio Pablo Cassará group for ARVAC, Agostina Demaría, Lineia Prado, Celeste Pueblas Castro, Lucas Saposnik, Jorge Geffner, Federico Montes de Oca, Julio C. Vega, Juan Fló, Pablo Bonvehí, Jorge Cassará, Karina A. Pasquevich, Juliana Cassataro

## Abstract

**Background:** This study (ARVAC-F2-3-002) assessed the immunogenicity, safety, and tolerability of a recombinant booster vaccine (ARVAC) containing the receptor binding domain of the SARS-CoV-2 Spike protein in three different versions: Gamma (ARVACGamma), Omicron BA.4/5 (ARVACOmicron), and Gamma/Omicron Bivalent (ARVACBivalent).

**Methods:** Randomized, double-blind, crossover, placebo-controlled, multicenter (11 centers in Argentina) Phase II/III trial including adult volunteers previously vaccinated against SARS-CoV-2 with ≤3 booster doses. Participants were randomized to receive ARVACGamma (50 µg)+placebo and vice-versa (1:1 ratio) (Phase II), and ARVACGamma (50 µg)+placebo, ARVACOmicron (50 µg)+placebo, and ARVACBivalent (Gamma/Omicron 25 µg/25 µg)+placebo and vice-versa (Phase III) (1:1:1:1:1:1 ratio) 28 days apart. The primary endpoint was the seroconversion rate of neutralizing antibodies compared to placebo. The vaccine immunogenicity was considered acceptable at >75% seroconversion rate to variants homologous to the antigen contained in the vaccine (prespecified primary endpoint).

**Results:** Participants (n=2012) (mean 48.2 years, SD 16.7; 48.1% women) were randomized and allocated to ARVACGamma (n=232 in Phase II and n=592 in Phase III), ARVACOmicron (n=594), and ARVACBivalent (n=594); 232 in Phase II and 370 in each Phase III group were included in the immunogenicity subset. Seroconversion rates to all SARS-CoV-2 variants were significantly higher after receiving any vaccine than placebo. All vaccine versions met the prespecified primary endpoint in all participants and in those 18−60 years old. In participants >60 years, the ARVACOmicron and the ARVACBivalent met the prespecified primary endpoint, whereas the ARVACGamma did not. The ARVACBivalent induced seroconversion rates were significantly higher than 75% across all tested SARS-CoV-2 variants (homologous and heterologous) and age groups. No vaccine-related serious adverse events were recorded; most local and systemic adverse events were grade 1-2.

**Conclusion:** Booster vaccination with Gamma, Omicron BA.4/5, and Bivalent protein subunit recombinant ARVAC vaccine versions elicited protective neutralizing antibody responses to several SARS-CoV-2 variants, with very low reactogenicity and a favorable safety profile.

Trial registration: NCT05752201

## Background

Coronavirus disease 2019 (COVID-19) continues to be a global health threat.^1,2^ Public health measures, population immunization, and the development of effective vaccines contributed to decreasing SARS-CoV-2 virus circulation, disease severity, and associated mortality.^3^ However, vaccine- and infection-induced immunity progressively wanes,^4,5^ and new, highly contagious SARS-CoV-2 variants and subvariants that escape from vaccine-induced immunity continue to emerge.^5,6^ In this scenario, primary vaccination schemes based on the ancestral SARS-CoV-2 variants fail to provide sufficient long-term protection.^4^

To ensure long-term immune memory, the WHO recommends homologous and heterologous booster doses after primary vaccination for protection against severe COVID-19 disease and death.^7^ Of the most common COVID-19 vaccine platforms, including inactivated viruses, viral vectors, RNA, and recombinant protein subunits, RNA vaccines are the most widely used. However, they are unstable and require storage at freezing temperatures (-20°C or -80⁰C), limiting their distribution, particularly in low- and middle-income countries.^5,8^ Conversely, recombinant subunit vaccines are more stable and may be stored in coolers, simplifying the storage and distribution logistics. Recombinant protein large-scale production is available, in several countries, enabling local manufacturing and widespread distribution with lower production costs.^9^ Despite the slower development speed of recombinant subunit vaccines compared to other vaccine platforms,^9^ they can also be modified to induce immunity against novel SARS-CoV-2 variants.^10^ Given that their safety profile record is well known and has been studied for more than 30 years, protein-based recombinant vaccines may be used in children, in elderly, and pregnant women.^11^

Argentina has developed and manufactured a recombinant protein subunit vaccine, ARVAC, which has been recently approved.^12^ The first version of the vaccine contains the receptor binding domain (RBD) of the spike protein of the SARS-CoV-2 Gamma variant, with K417T, E484K, and N501Y mutations. Preclinical studies demonstrated that the Gamma RBD version is more immunogenic than the ancestral RBD at inducing broader neutralizing antibodies (nAbs) even against distant variants, such as the Omicron BA.5.^13^ In a Phase I trial, the vaccine was safe and elicited a robust and broad nAb response against several SARS-CoV-2 variants.^14^

In this work, we present the results of a Phase II/III trial. The study is a randomized, placebo-controlled trial assessing the immunogenicity, safety, and tolerability of Gamma, Omicron BA.4/5, and bivalent versions of the ARVAC vaccine, used as a booster in adult volunteers previously immunized with different SARS-CoV-2 vaccine platforms.

## Methods

### Study Design, Objectives, Participants, and Oversight

The ARVAC-F2-3-002 study is a multicenter, randomized, double-blind, crossover, placebo-controlled Phase II/III trial evaluating the immunogenicity, safety, and tolerability of a recombinant protein vaccine against SARS-CoV-2 in adult (≥18 years) volunteers previously vaccinated against SARS-CoV-2 with ≤3 booster doses. Additional inclusion and exclusion criteria are provided in the **Appendix**. Investigators from 11 participating centers in Argentina (listed in the **Appendix**) consecutively recruited volunteers.

The “Centro de Educación Médica e Investigaciones Clínicas – CEMIC” (Buenos Aires, Argentina) is the sponsor. An external, independent data safety monitoring board reviewed safety data. The trial adhered to the International Council for Harmonization of Technical Requirements for Pharmaceuticals for Human Use (ICH) Guideline for Good Clinical Practice (GCP) and the local data protection law (“Ley de protección de datos” 25,326). All participants signed the informed consent form. The National Administration of Drugs, Food, and Medical Technology (*Administración Nacional de Medicamentos, Alimentación y Tecnología Médica,* ANMAT), the CABA Local Ethics Committee (PRIISA, *Plataforma de Registro Informatizado de Investigaciones en Salud de Buenos Aires*), and the Ethics Committee on Clinical Research (CEIC) of the Infectious Studies Center S.A. (*Centro de Estudios Infectológicos,* CEI) - Stamboulian approved the protocol, which was prospectively registered at ANMAT, PRIISA, and clinicaltrials.gov (NCT05752201). The local Ethics Committees that approved the study protocol are listed in the **Appendix**.

### Recombinant Protein Vaccines

The vaccine antigen encompassed aminoacids 319R-537K in the RBD of the SARS-CoV- 2 Spike protein. Recombinant proteins for the variants Gamma and Omicron BA.4/5 were produced in CHO-S cells.^14^ The Laboratorio Pablo Cassará S.R.L. (Buenos Aires, Argentina) manufactured the ARVAC vaccine as a liquid suspension formulation containing 50 µg of recombinant protein in a 0.5 mL vial, adjuvanted with aluminum hydroxide gel (alhydrogel, 0.5 mg).

### Randomization and Procedures

Participants were recruited in two stages. In stage 1 (Phase II), participants were randomized into two subgroups at a 1:1 ratio to receive Gamma-based vaccine (ARVACGamma) (50 µg) + placebo (group A) and placebo + ARVACGamma (50 µg) (group B) 28 days apart. In stage 2 (Phase III), participants were randomized into three groups to receive the ARVACGamma (50 µg), the Omicron BA.4/5-based (ARVACOmicron) (50 µg), and the Bivalent (Gamma/Omicron BA.4/5 25 µg/25 µg, ARVACBivalent) vaccine, with two subgroups each, receiving vaccine + placebo (group A) or placebo + vaccine (group B) 28 days apart (1:1:1:1:1:1 ratio) (Figure S1). Within each group, individuals were assigned to age subgroups (18‒60 years and >60 years) and to immunogenicity subsets (**Appendix**).

Assessments were performed during five visits (V): V1 on day 1 (inclusion visit); V2, 14 ±2 days later; V3, 28 ±2 days after V1; V4, 56 ±2 days after V1; and V5, 90 ±2 days after V1. In all groups, the first treatment was administered on V1 and the second on V3.

### Immunogenicity Endpoints and Variables

NAbs against Ancestral (Wuhan), Gamma, or Omicron BA.5 SARS-CoV-2 variants were measured on plasma samples obtained before and after first treatment on days 1 (d1) and 14 (d14) respectively and presented as geometric mean titers (GMT); geometric mean fold rises (GMFR) and GMT ratios (GMTR) were calculated. Additionally, titers against the SARS-CoV-2 Ancestral (Wuhan) variant were transformed to IU/mL using a secondary standard calibrated with a WHO international standard.^15^ Based on previous studies, a >1030 UI/mL threshold of nAbs was associated with a 90% efficacy against symptomatic infection.^16^

The study’s primary endpoint was the seroconversion rate 14 days after receiving the vaccine compared to placebo. The vaccine immunogenicity was considered acceptable at seroconversion rate >75% to variants homologous to the antigen contained in the vaccine (prespecified primary endpoint). The threshold for seroconversion was defined as a 4- fold or a 2-fold increase in nAb titers for individuals with “low” or “high” baseline nAb levels against Ancestral SARS-CoV-2 (<949 or ≥949 IU/mL), respectively. This classification considered a lower nAb increment in individuals with high basal titers and the association of nAb levels ≥949 IU/mL with high vaccine protection against symptomatic infection.^16^ In Phase III, seroconversion rates were analyzed according to age group and vaccine version.

Other immunogenicity endpoints were the comparison of seroconversion rates among vaccine variants and the superiority and inferiority of bivalent vs. monovalent vaccines. Additional secondary and exploratory endpoints and methods are described in the **Appendix**.

### Safety Endpoints and Assessments

Safety endpoints were solicited local and systemic adverse events (AEs), registered daily in the participants’ diary within seven days after each vaccine dose, and unsolicited local AEs occurring within 20 minutes after administration. AEs were classified according to severity and their relationship to the study medication based on published guidelines.^17^ AEs and serious AEs (SAEs) occurring from the first administration until one and six months after the last administration, respectively, were recorded. Additional details are provided in the **Appendix**.

### Statistical Analysis

The sample size was calculated based on the ARVACGamma vaccine seroconversion rates obtained in a previous Phase I study.^14^ For the prespecified primary endpoint, the estimated sample size was 113 participants for each vaccine candidate, considering a 10% dropout rate. To assess the exploratory endpoint of seroconversion superiority of the bivalent vs. the monovalent vaccines, a sample size of 248 participants for each vaccine candidate was calculated (276, considering a 10% dropout rate). For the safety analysis, a sample size of 2014 participants, 232 in Phase II and 1782 in Phase III, was estimated to detect AEs with a 0.1% prevalence, considering a 20% dropout rate. The Appendix includes a detailed description of the sample size calculation.

The primary endpoint (i.e., immunogenicity) was evaluated in the population of participants randomized to immunogenicity subset who received at least one dose of vaccine or placebo using samples for determination of total and nAbs before (d1) and after the first administration (d14) within an appropriate time frame. The safety endpoints were evaluated in all participants who received at least one dose of vaccine or placebo.

Seroconversion rates after vaccine administration were compared to placebo using the Fisher’s exact test and to the 75% reference using a Z-test (primary endpoint). NAb titers were compared between vaccine variants using the Kruskal-Wallis and the Dunns tests for multiple comparisons, and between timepoints (d1 vs. d14) using the Wilcoxon test for paired data. Additional statistical methods are included in the **Appendix**. Significance was set at a bilateral α<0.05. Statistical analyses were performed using GraphPad Prism v8.4.2 (GraphPad Software, San Diego, CA) or R version 4.3.2 for MacOS (R Foundation for Statistical Computing, Viena, Austria).

## Results

### Participants

A total of 2126 volunteers signed the informed consent, 2012 were included (232 in Phase II and 1780 in Phase III) and randomized. All received the first vaccine/placebo administration and 1905 the second; 138 discontinued the study, and 1874 finished the study protocol (Figure 1).

**Figure 1.**
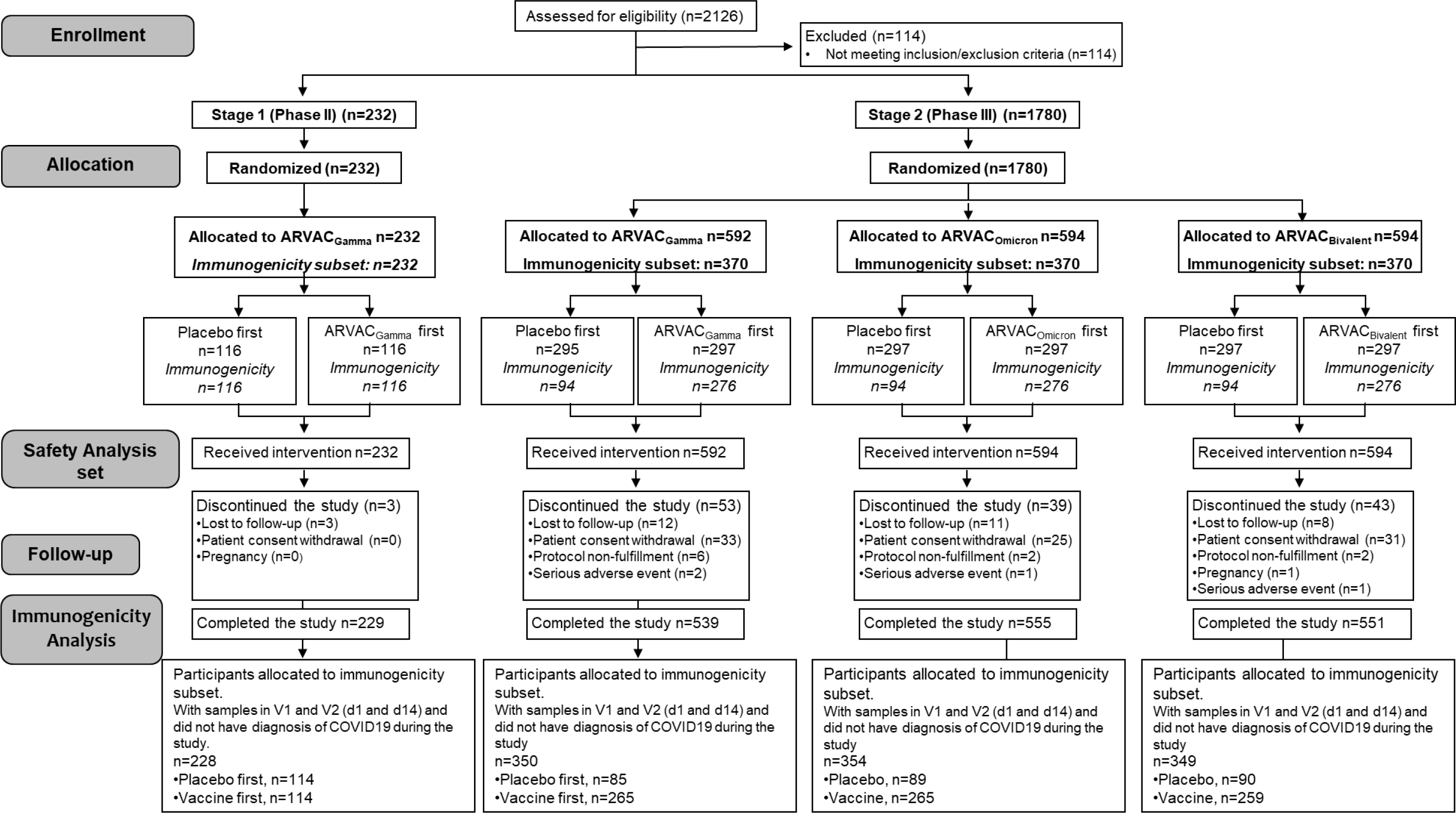
Flow chart of study participants.

The mean (SD) age was 48.2 (16.7) years; 48.1% were women, 64.7% had previously received a complete vaccination scheme with one booster, and 44.2% had previous diagnostic of COVID-19 (Table 1). The immunogenicity subset included all study participants of Phase II (Table 1) and 1110 participants of Phase III (Table S1).

**Table 1.** Characteristics of study participants according to study Phase and sex. N=2012.

|  | Phase II |  |  | Phase III |  |  | All |  |  |
| --- | --- | --- | --- | --- | --- | --- | --- | --- | --- |
|  | Female | Male | Total | Female | Male | Total | Female | Male | Total |
| N (%) | 134 (57.8) | 98 (42.2) | 232 | 834 (46.9) | 946 (53.1) | 1780 | 968 (48.1) | 1044 (51.9) | 2012 (100) |
| Age (years), mean (SD) | 36.6 (12.1) | 35.7 (11.5) | 36.2 (11.8) | 47.3 (16.7) | 52.0 (16.3) | 49.8 (16.6) | 45.8 (16.6) | 50.5 (16.6) | 48.2 (16.7) |
| BMI (kg/m<sup>2</sup>), mean (SD) | 27.0 (4.8) | 26.9 (4.1) | 27.0 (4.5) | 26.7 (4.6) | 27.4 (3.9) | 27.1 (4.2) | 26.8 (4.6) | 27.3 (3.9) | 27.1 (4.3) |
| Number of boosters after complete primary vaccination scheme, n (%) |  |  |  |  |  |  |  |  |  |
| 0 | 46 (34.3) | 40 (40.8) | 86 (37.1) | 93 (11.2) | 113 (19.9) | 206 (11.6) | 139 (14.4) | 153 (14.7) | 293 (14.5) |
| 1 | 88 (65.7) | 58 (59.2) | 146 (62.9) | 538 (64.5) | 526 (55.6) | 1064 (59.8) | 626 (64.7) | 584 (55.9) | 1210 (60.1) |
| 2 |  |  |  | 101 (12.1) | 166 (17.5) | 267 (15.0) | 101 (10.4) | 166 (15.9) | 267 (13.3) |
| 3 |  |  |  | 102 (12.2) | 141 (14.9) | 243 (13.6) | 102 (10.5) | 141 (13.5) | 243 (12.1) |
| Time since last vaccination (months), |  |  |  |  |  |  |  |  |  |
| mean (SD) | 13.2 (2.7) | 13.1 (3.2) | 13.2 (2.9) | 14.8 (4.1) | 14.4 (4.6) | 14.3 (4.4) | 14.3 (4.3) | 14.3 (4.5) | 14.1 (4.3) |
| Previous COVID-19, n (%) |  |  |  |  |  |  |  |  |  |
| No | 87 (64.9) | 73 (74.5) | 160 (69.0) | 406 (48.7) | 557 (58.9) | 963 (54.1) | 493 (50.9) | 630 (60.3) | 1123 (55.8) |
| Yes | 47 (35.1) | 25 (25.5) | 72 (31.0) | 428 (51.3) | 389 (41.1) | 817 (45.9) | 475 (49.1) | 414 (39.7) | 889 (44.2) |
| Time since infection (months), |  |  |  |  |  |  |  |  |  |
| mean (SD) | 20.4 (8.1) | 20.4 (9.6) | 20.4 (8.6) | 19.8 (9.0) | 20.8 (8.6) | 20.0 (8.7) | 19.8 (9.0) | 20.8 (8.6) | 20.0 (8.7) |
| Study treatment, n (%) |  |  |  |  |  |  |  |  |  |
| Bivalent |  |  |  | 139 (16.7) | 158 (16.7) | 297 (16.7) | 139 (14.3) | 158 (15.1) | 297 (14.8) |
| Gamma | 70 (52.2) | 46 (46.9) | 116 (50.0) | 136 (16.3) | 161 (17.1) | 297 (16.7) | 206 (21.3) | 207 (19.9) | 413 (20.5) |
| Omicron |  |  |  | 144 (17.2) | 153 (16.2) | 297 (16.7) | 144 (14.9) | 153 (14.6) | 297 (14.8) |
| Placebo | 64 (47.8) | 52 (53.1) | 116 (50.0) | 415 (49.8) | 474 (50.1) | 889 (49.9) | 479 (49.5) | 526 (50.4) | 1005 (50.0) |

BMI, body mass index; SD, standard deviation.

### Seroconversion Rates (Primary Endpoint)

Seroconversion rates to homologous and non-homologous SARS-CoV-2 variants were higher after receiving any vaccine than placebo overall and in the two age groups (*p*<0.0001 for all comparisons) (Table 2). All vaccine versions met the prespecified primary endpoint (i.e., seroconversion rate >75% reference for the homologous variant) in all participants and in those 18−60 years old. In participants >60 years, the ARVACOmicron and the ARVACBivalent met the prespecified primary endpoint, whereas the ARVACGamma did not. The ARVACBivalent induced seroconversion rates were significantly higher than 75% across all tested SARS-CoV-2 variants (homologous and heterologous) and age groups (*p*<0.001). Analyses using normalized antibody titers yielded similar results (Table S2).

**Table 2.**
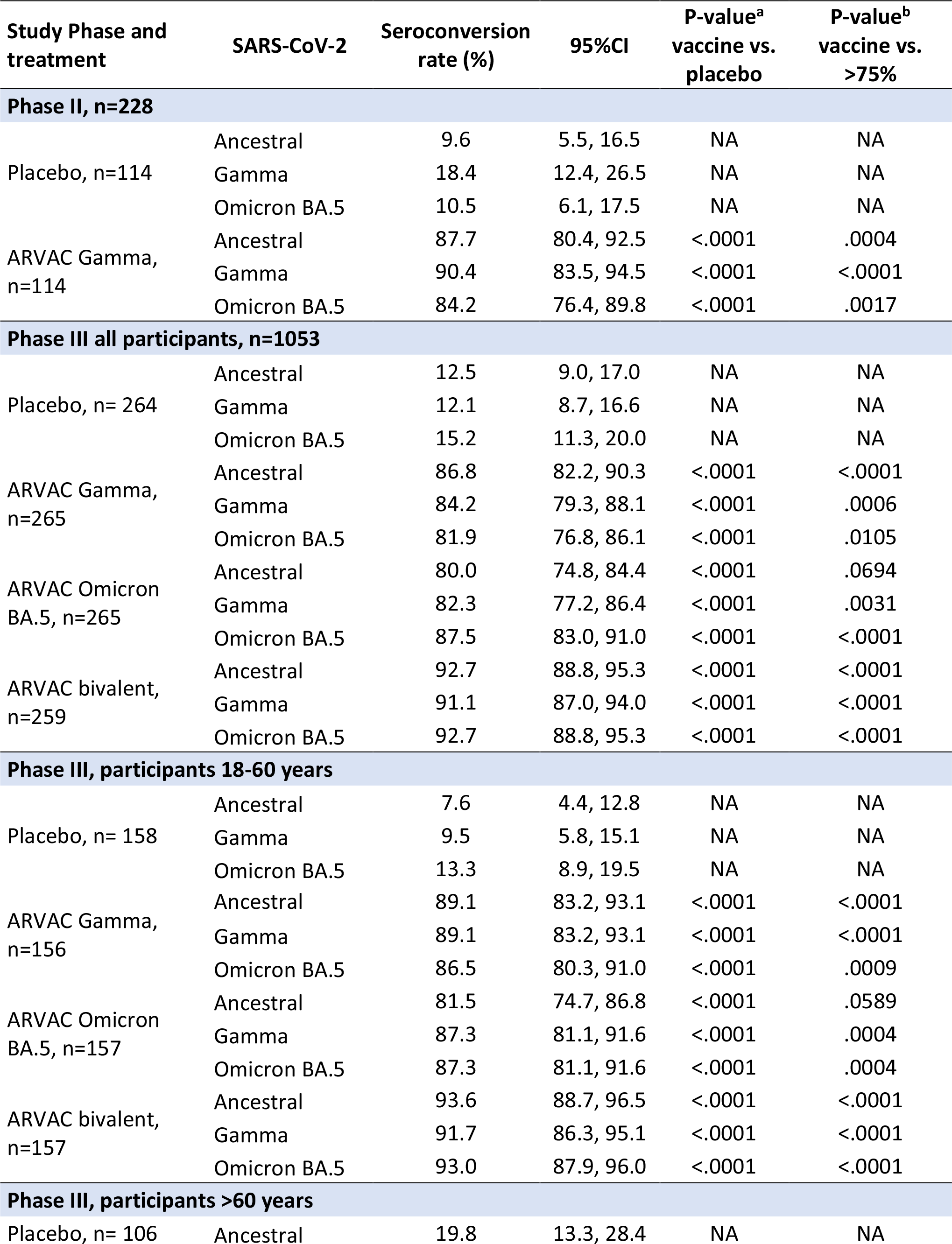
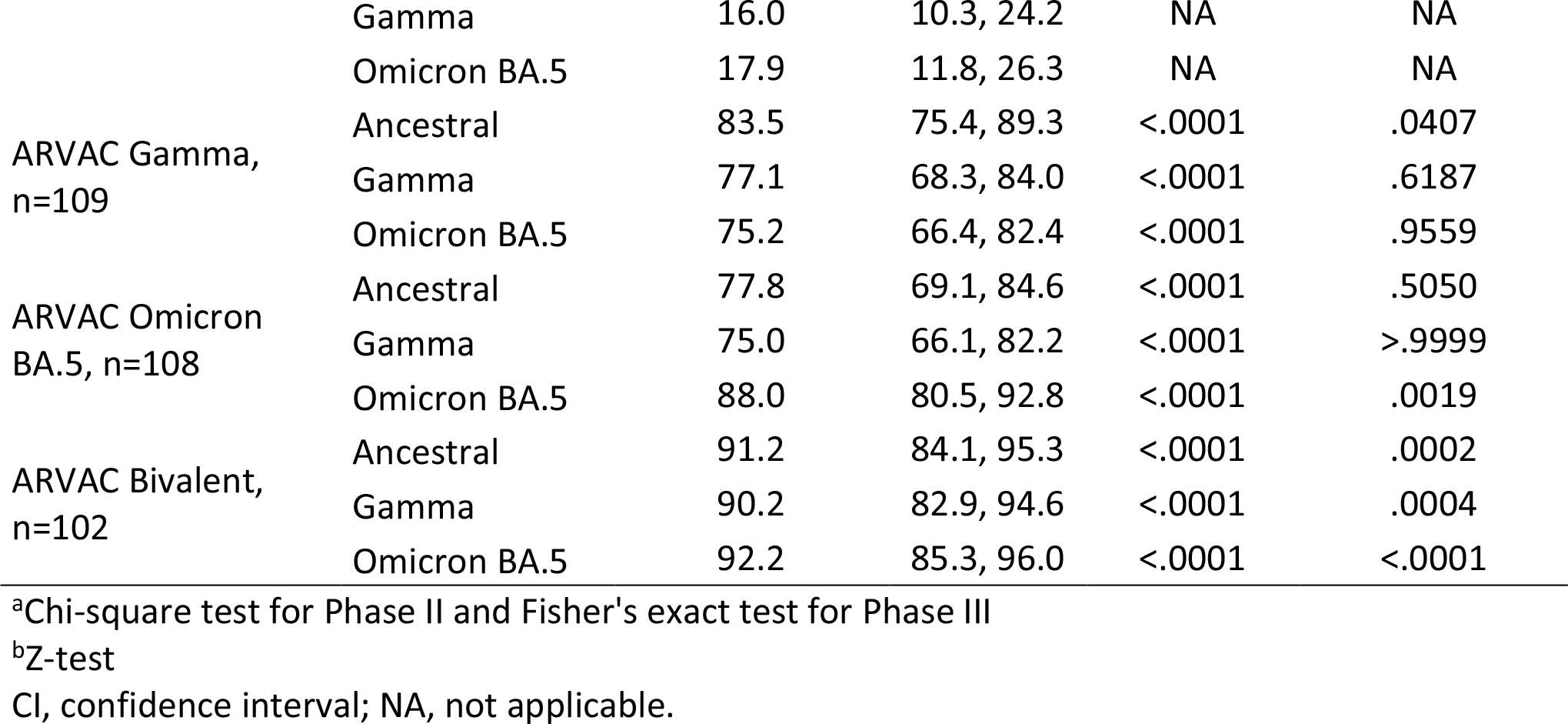
Seroconversion rates in Phase II (n=228) and Phase III (n=1053) participants for the different vaccine variants and age groups compared to placebo and a >75% reference.

### Neutralizing Antibody Titers (Secondary Endpoints)

GMTs to Ancestral, Gamma, and Omicron variants increased from d1 to d14 in participants receiving any vaccine (*p*<0.0001 for all comparisons) but not in those receiving placebo overall and in the two age groups (Figure 2). The percentages of participants with >8 GMFR from d1 to d14 are summarized in Table S3.

**Figure 2.**
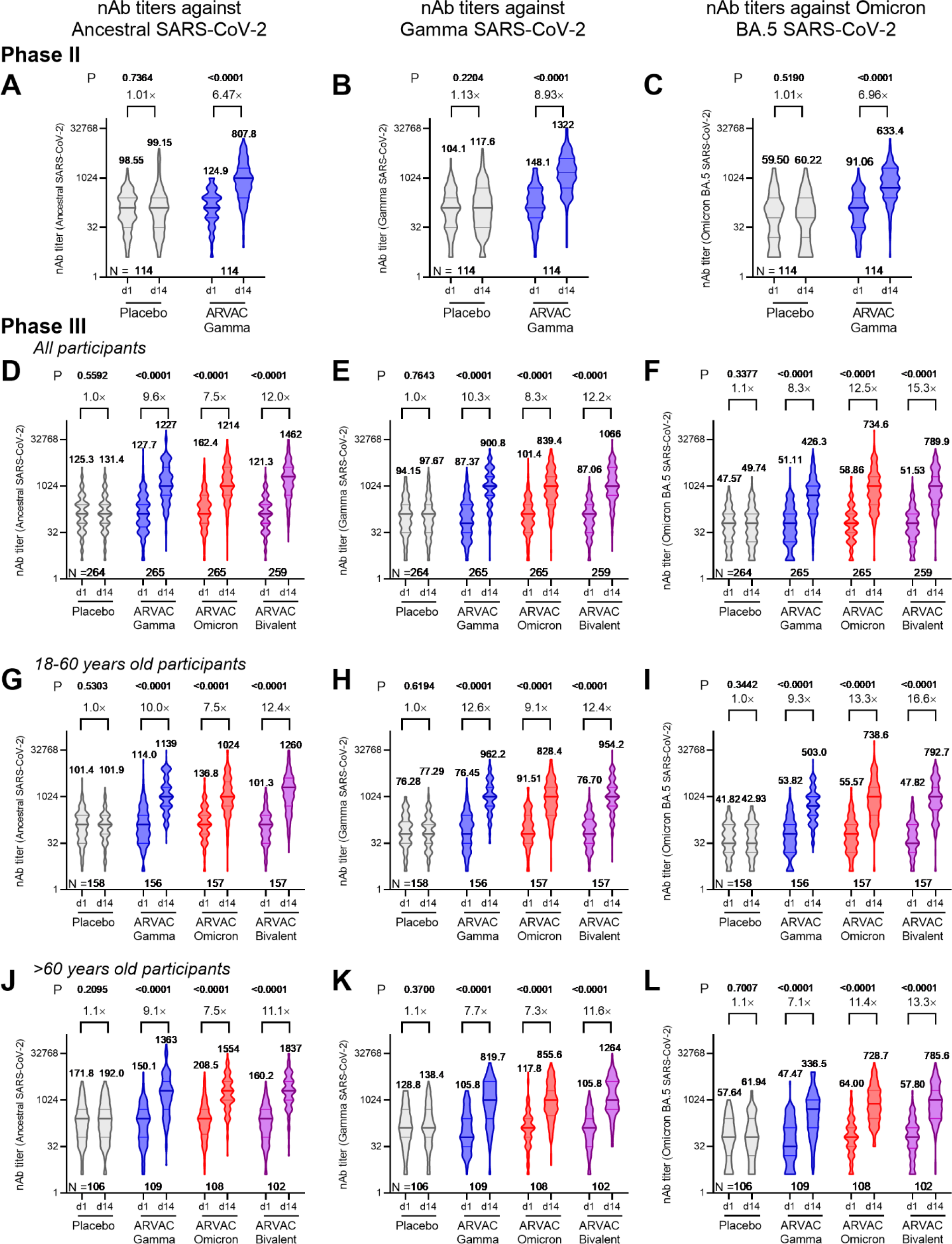
Neutralizing antibody titers against SARS-CoV-2 variants before (d1) and 14 days after vaccine/placebo administration. The plots represent antibody titers against SARS-CoV-2 Ancestral (A, D, G, J), Gamma (B, E, H, K), and Omicron variants (C, F, I, L) in plasma samples of Phase II (A-C) and Phase III (D-F) participants. Phase III participants were classified according to age into 18-60 years (G-I) and >60 years (J-L). The thick horizontal lines in the violin plots represent median. Geometric mean titers (GMTs) value is indicated above the plots. Geometric mean fold rises (GMFR) and *p*-values comparing titers before and after administration are indicated. *P*-values were calculated using the non-parametric paired Wilcoxon test. nAb, neutralizing antibodies.

At day 90 (d90), GMFR remained statistically significant for all vaccine versions and SARS-CoV-2 variants and according to age group (Figures S2) (*p*<0.0001 for all comparisons).

Participants with nAbs to the Ancestral variant >1030 UI/mL increased at d14 for all vaccine versions (*p*<0.0001), with similar results in the two age groups. For ARVACGamma, ARVACOmicron, and ARVACBivalent vaccines, percentages raised from 30.8%, 31.2%, and 24.8% to 87.2%, 85.4%, and 87.9%, respectively in participants 18−60 years; in participants >60 years from 40.4%, 47.2%, and 45.1% to 84.4%, 89.8%, and 92.2%, respectively (Figure S3).

### Analyses According to Previous Vaccination and COVID-19 Infection (Secondary Endpoints)

GMTs to all SARS-CoV-2 variants increased in participants receiving the ARVAC vaccine regardless of the number of previous booster doses (Figure S4) or the primary vaccine platforms (adenovirus, mRNA, inactivated virus, heterologous vaccination, or virus like particles) (Figures S5-S8). GMTs and GMFRs lacked significant differences according to previous COVID-19 infection (Figure S9).

### Seroconversion Rates and nAb Titers According to Vaccine Versions (Exploratory Endpoints)

ARVACBivalent was not inferior to ARVACOmicron and ARVACGamma regarding seroconversion rates against the three SARS-CoV-2 variants in all participants and according to age groups (*p*<0.001) (Table S4). ARVACBivalent seroconversion rates were superior to ARVACGamma’s against the Omicron variant (*p*=0.001) and to ARVACOmicron’s against the Ancestral (*p*<0.001) and Gamma (*p*=0.013) variants. In participants aged 18−60 years, ARVACBivalent seroconversion rates were superior to ARVACOmicron’s against the Ancestral variant (*p*=0.006), and in participants >60 years, they were superior to ARVACGamma‘s against the Omicron variant (*p*=0.04) and to ARVACOmicron’s against the Ancestral and Gamma variants (*p*=0.020) (Table S5).

An adjusted multivariate analysis confirmed the superiority of ARVACBivalent to ARVACGamma in seroconversion rates against the Ancestral (*p*=0.030), Gamma (*p*=0.015), and Omicron (*p*<0.001) variants and to ARVACOmicron against the Ancestral (*p*<0.001) and Gamma (*p*=0.005) variants (Tables S6-S8).

Accordingly, the ARVACBivalent was not inferior to the ARVACOmicron and the ARVACGamma regarding GMT against the three SARS-CoV-2 variants regardless of age (*p*<0.001) (Table S9). All vaccine versions induced similar GMTs to the Ancestral and Gamma variants. However, either in all participants or in those >60 years, the ARVACBivalent (p=0.0002 and p=0.0026, respectively) and the ARVACOmicron (p=0.0033 and p=0.0105, respectively) induced higher titers against the Omicron variant than the ARVACGamma (Tables S10-S12).

The ARVACBivalent induced higher GMFRs than the ARVACOmicron for Ancestral (p=0.0014) and Gamma (p=0.0076) variants and higher than the ARVACGamma for the Omicron variant (p=0.0001) (Table S13).

GMTR analysis showed that the ARVACBivalent was superior to the ARVACGamma in nAbs responses to Gamma (p=0.048) and to Omicron (p<0.001) variants. The ARVACBivalent was superior to the ARVACOmicron in the nAb response to Ancestral (p=0.022) and to Gamma (p=0.013) variants (Table S14).

### Anti-Spike-Specific Antibodies and Mucosal Response (Exploratory Endpoints)

Plasma levels of anti-spike-specific IgG increased (d1 to d14) in participants receiving any vaccine, regardless of age group (*p*<0.0001) (Figures S10); changes remained significant at d90 (*p*<0.0001) (Figure S11). Anti-S1-specific IgA in saliva increased in participants receiving any vaccine (*p*<0.0001) (Figure S12).

### NAbs Against New Emerging Virus Variants

The ARVACBivalent vaccine booster activity was further studied against new predominant Omicron subvariants that circulated recently in Argentina. GMTs to XBB.1.18 and JN.1 subvariants increased significantly in participants 18-60 years (*p*<0.0001 and *p*=0.0015, respectively) and in participants >60 years (*p*=0.0001 and *p*=0.0069, respectively) (Figure S13,). While most participants (>89%) had detectable nAbs titers against Ancestral, Gamma, and Omicron BA.5 variants before vaccination, nAbs to XBB.1.18 and JN.1 were detectable in 50.0% and 18.8% of participants aged 18-60 and in 61.5% and 33.0% of those >60 years, respectively. These percentages increased to 91.7% and 83.3% in participants aged 18-60 and to 100% and 92.3% in participants >60 years after ARVACBivalent administration (Figure S13).

### Safety

Most local and systemic AEs were Grade 1 and 2 (Table 3), and no SAEs related to the vaccine were reported. The most frequent local AEs were pain and sensitivity/discomfort in the injection site and were more frequent in participants receiving the vaccine than placebo (*p*<0.001) (Table 3). Pain was more frequent after administration of the ARVACOmicron and ARVACBivalent versions (*p*<0.001) (Table S15).

**Table 3.** Local and systemic adverse reactions according to severity and treatment (vaccine vs. placebo), n (%). N=2012.

|  | Vaccine (n=1960) |  |  |  |  | Placebo (n=1957) |  |  |  | P-value** |
| --- | --- | --- | --- | --- | --- | --- | --- | --- | --- | --- |
|  | Grade 1 | Grade 2 | Grade 3 | Grade 4 | Total* | Grade 1 | Grade 2 | Grade 3 | Total* |  |
| Local, n (%)# |  |  |  |  |  |  |  |  |  |  |
| Pain | 810 (96.3) | 31 (3.7) | 0 | 0 | 841 (42.9) | 601 (98.0) | 11 (1.8) | 1 (0.2) | 613 (31.3) | <.001 |
| Sensitivity/discomfort | 565 (88.6) | 69 (10.8) | 4 (0.6) | 0 | 638 (32.5) | 385 (92.1) | 30 (7.2) | 2 (0.5) | 417 (21.3) | <.001 |
| Swelling/induration | 155 (98.1) | 3 (1.9) | 0 | 0 | 158 (8.1) | 73 (100) | 0 | 0 | 73 (3.8) | .257 |
| Erythema/redness | 74 (96.1) | 3 (3.9) | 0 | 0 | 77 (3.9) | 44 (97.8) | 1 (2.2) | 0 | 45 (2.3) | .660 |
| Itching | 54 (100) | 0 | 0 | 0 | 54 (2.8) | 28 (96.6) | 1 (3.4) | 0 | 29 (1.5) | .995 |
| Systemic, n (%)# |  |  |  |  |  |  |  |  |  |  |
| Diarrhea | 43 (91.5) | 3 (6.4) | 1 (2.1) | 0 | 47 (2.4) | 38 (97.4) | 1 (2.6) | 0 | 39 (2.0) | .388 |
| Headache | 203 (83.9) | 37 (15.3) | 2 (0.8) | 0 | 242 (12.3) | 170 (90.4) | 17 (9.0) | 1 (0.5) | 188 (9.6) | .006 |
| Joint pain | 48 (85.7) | 7 (12.5) | 1 (1.8) | 0 | 56 (2.9) | 39 (84.8) | 7 (15.2) | 0 | 46 (2.4) | .321 |
| Muscle pain/myalgia | 103 (86.6) | 15 (12.6) | 1 (0.8) | 0 | 119 (6.1) | 93 (86.9) | 14 (13.1) | 0 | 107 (5.5) | .420 |
| Chills | 34 (81.0) | 7 (16.7) | 1 (2.4) | 0 | 42 (2.1) | 26 (89.7) | 3 (10.3) | 0 | 29 (1.5) | .122 |
| Fatigue/tiredness/decay | 206 (88.8) | 23 (9.9) | 2 (0.9) | 1 (0.4) | 232 (11.8) | 169 (89.9) | 19 (10.1) | 0 | 188 (9.6) | .024 |
| Fever | 22 (75.9) | 6 (20.7) | 1 (3.4) | 0 | 29 (1.5) | 14 (93.3) | 1 (6.7) | 0 | 15 (0.8) | .034 |
| Nausea | 32 (94.1) | 2 (5.9) | 0 | 0 | 34 (1.7) | 31 (91.2) | 3 (8.8) | 0 | 34 (1.7) | .993 |
| Palpitations | 15 (83.3) | 3 (16.7) | 0 | 0 | 18 (0.9) | 12 (92.3) | 1 (7.7) | 0 | 13 (0.7) | .370 |
| Drowsiness | 196 (88.3) | 24 (10.8) | 2 (0.9) | 0 | 222 (11.3) | 183 (92.0) | 16 (8.0) | 0 | 199 (10.2) | .243 |
| Vomiting | 6 (100) | 0 | 0 | 0 | 6 (0.3) | 3 (50.0) | 3 (50.0) | 0 | 6 (0.3) | .997 |

Adverse reactions to all vaccine and placebo administrations are included.

The most frequent systemic AEs were headache and fatigue/tiredness/decay in 12.3% and 11.8% of participants receiving the vaccine respectively, and 9.6% in participants receiving the placebo (Table 3). A description of AEs according to the vaccine version is included in Table S16.

## Discussion

This randomized, double-blind, crossover, placebo-controlled Phase II/III trial showed that booster vaccination with Gamma, Omicron BA.4/5, and bivalent versions of the protein subunit recombinant ARVAC vaccine elicited robust antibody responses to SARS-CoV-2 Ancestral, Gamma, and Omicron BA.5 variants in adults, regardless of primary vaccination platform, and previous SARS-CoV-2 infection. At d14 after vaccination, seroconversion rates to homologous and non-homologous SARS-CoV-2 variants after receiving any vaccine version were higher than placebo. In all participants the three vaccine versions elicited seroconversion rates to homologous SARS-CoV-2 variants >75% reference (prespecified primary endpoint). GMTs against the three SARS-CoV-2 variants significantly increased, and antibody responses persisted for at least 90 days, even in participants >60 years. The nAb levels in IU/mL suggested that the vaccine versions achieved ≥90% efficacy against symptomatic infection in 84.4%‒92.2% of participants. The bivalent vaccine induced more prominent GMFRs and higher seroconversion rates than monovalent vaccines. Moreover, all vaccine versions increased anti-spike-specific antibody levels in plasma and saliva, with a favorable safety and tolerability profile.

Results from this trial confirmed those from our previous Phase I study, including younger participants (18-55 years) with a variety of primary vaccination schemes.^14^ Moreover, the results from this trial support previous studies indicating increased immunogenicity and breadth of a Gamma-variant vaccine compared to the ancestral- variant vaccine.^13,14^

To our knowledge, few trials have simultaneously assessed and compared the immunogenicity outcomes of several vaccine variants, as showed in this Phase III trial, comparing three vaccine versions.^18,19^ Bivalent Ancestral/Omicron, Alpha/Beta, and Ancestral/Beta recombinant boosters have previously shown robust nAb responses in individuals who had received primary schemes based mostly on mRNA or Adenoviruses,^18–20^ but a bivalent recombinant booster vaccine lacking the Ancestral/Alpha variants remained unassessed. Despite including a lower dose of each monovalent vaccine, the ARVACBivalent booster was non-inferior regarding seroconversion rates and GMTs, and, remarkably, it was superior to the monovalent vaccines against heterologous SARS-CoV-2 variants. Furthermore, ARVACBivalent induced seroconversion rates >90% against all SARS-CoV-2 variants in all age groups. Similar results, superiority to monovalent versions against heterologous variants and non-inferiority against homologous variants, were described for a bivalent Omicron BA.5/ancestral SARS-CoV-2 recombinant spike protein vaccine as a heterologous booster dose.^19^

Unlike this trial, the population included in other trials assessing boosting with recombinant protein vaccines were highly homogenous regarding the primary vaccination scheme, mostly based on mRNA (BNT162b2 or mRNA-1273), Ad26, and ChAdOx-1 vaccines.^18–21^ However, at least seven vaccines based on different platforms were applied as primary and booster doses in Argentina when we initiated this study.^22^ This trial showed robust nAb responses regardless of the previous vaccination scheme (six platforms) and the number of previous booster doses (no booster, one, or two). Hence, the results of this trial were obtained in a large population with no strict selection criteria, reflecting the variety of primary vaccination schemes in Argentina and providing valuable information for applying the ARVAC vaccine in real-world populations. In this regard, an adjusted multivariate analysis revealed the superiority of ARVACBivalent compared to ARVACGamma or ARVACOmicron independently of age, sex, number of previous vaccine doses, previous vaccine platform, time since last vaccination, and previous COVID-19 history. Moreover, these results contribute to the increasing evidence that heterologous schedules may provide superior immunogenicity to homologous booster schedules.^23–25^ Furthermore, this study included participants with and without comorbidities. The WHO includes older individuals (>50-60 years, depending on the country) as high-priority, with booster doses recommended at 12-month intervals.^7^ Hence, our results provide data on booster responses in relevant populations.

The Omicron BA.4/5 and Gamma antigens in the ARVACBivalent contain the main mutations conferring immune evasion and are, therefore, likely to elicit the production of antibodies neutralizing other Omicron subvariants. The immunogenicity of ARVACBivalent against Omicron XBB.1.18 and JN.1 subvariants is encouraging, given that these variants contain more immune-evasive mutations than most others detected to date and are predominant in many geographical centers. Nevertheless, it is possible that adapting ARVAC to new emerging variants (i.e., XBB.1.5 or JN.1) would further increase the immunogenicity. Nevertheless, the Omicron BA.4/5 is adapted to the currently circulating SARS-CoV-2 variants (i.e., Omicron subvariants).^26^ The WHO and the Food and Drug Administration (FDA) recommend that booster doses include an Omicron BA.4/5 component and exclude the Ancestral strain;^27,28^ therefore, the ARVAC vaccine assessed in this study fulfills these recommendations.

ARVAC demonstrated to be safe and as expected for being a protein-based recombinant vaccine induced a very low reactogenic response. Therefore, an alternative vaccine platform with a history of safe and effective use has the potential to benefit public health by providing an additional choice to our region.

One limitation of this study is the short follow-up for immunogenicity. However, in the Phase I study, the nAb titers remained significantly increased even after six ^14^ and twelve months of vaccine administration (unpublished results). Despite this limitation, this study showed that the ARVAC RBD-based protein vaccine and, particularly, its bivalent version, used as a first, second or third booster, elicited robust, protective, long-lasting antibody responses in participants with a complete vaccination scheme regardless of age, sex, previous vaccine platform, and previous SARS-CoV-2 infection.

## Conclusions

Booster vaccination with Gamma, Omicron BA.4/5, and bivalent versions of the protein subunit recombinant ARVAC vaccine elicited protective neutralizing antibody responses to several SARS-CoV-2 variants, including the currently circulating Omicron variant. ARVAC showed very low reactogenicity and a favorable safety profile, as expected for a recombinant protein alhydrogel adjuvanted vaccine. The ARVAC vaccine is a valuable booster option given the recommended administration of booster doses that include Omicron while excluding Ancestral SARS-CoV-2 variants to high-priority populations, the variety of approved primary vaccination schemes, the feasibility and low cost of large-scale recombinant vaccine production, its potential for adaptation, its safety profile, and its feasible widespread distribution.

## Declarations

### Availability of data and materials

The datasets generated and/or analyzed during the current study are available from the corresponding author on reasonable request.

## Acknowledgments

We thank all the trial participants, the staff members at each site for their high degree of professionalism in the conduct of the trial. We also thank the i2e3 Procomms team (Barcelona, Spain) and especially Sara Cervantes, Ph.D., for providing medical writing support during the manuscript preparation. We thank the administrative staff at University of San Martin and at INBIRS. We thank the support of the University of San Martin (UNSAM), the National Scientific and Technical Research Council (CONICET), National Ministry of Science and technology, the National Agency for promotion of Science and Technology of Argentina, the National Ministry of Health, the Ministry of Health of the Province of Buenos Aires and the National Administration of Drugs, Food and Medical Devices (ANMAT). We are thankful to the staff of the Pablo Cassará Laboratory for their essential contribution to the development, scaling, manufacturing, control, and stability studies of the clinical batches of antigens and vaccines. We thank Ethel Feleder and Karina E. Halabe from Clinical Pharma S.R.L. We thank Javier Mariarini from Hospital el Cruce -Florencio Varela, Buenos Aires, Argentina- and Fundación Huésped -CABA, Argentina- for statistical analysis. We are grateful to Dr. Ángela Spanguolo de Gentile, Dr. Roberto Debbag, and Dr. Lucas Giménez who were members of the external Independent Committee of Data Review for their dedication and continuous invaluable advice.

**Laboratorio Pablo Cassará Group for ARVAC:** Sabrina A. Del Priore; Andrés C. Hernando Insua; Ingrid G Kaufman; Adrián Di María; Adrián Gongora; Agustin Moreno; Susana Cervellini; Martin Blasco; Fernando Toneguzzo.

