## Appendix. Supplementary Methods, Tables and Figures for "Immunogenicity and Safety of Gamma, Omicron BA.4/5 and Bivalent SARS-CoV-2 RBD-based Protein Booster Vaccines in Adults Previously Immunized with Different Vaccine Platforms: a Phase II/III, Randomized, Clinical Trial"

#### *Inclusion and exclusion criteria*

##### *Inclusion criteria*

Volunteers had to be negative for SARS-CoV-2 in a polymerase chain reaction or antigen test at study inclusion. Volunteers were required to have the ability and willingness to comply with the prohibitions and restrictions specified in the protocol, the ability to read, comprehend, and complete the electronic questionnaires about COVID-19 symptoms and signs, and the ability to provide informed consent. Volunteers had to accept the prohibition to donate bone marrow, blood, or blood derivatives until 3 months after the last vaccine administration. Fertile female volunteers were required to have a negative pregnancy test at study initiation and commitment to use contraception from the informed consent signature until 3 months after the vaccine administration. Hormonal contraceptive methods were required to start at least 28 days before the first vaccine administration.

In Stage I (Phase II), volunteers were required to be 18 to 60 years old with no known comorbidities. In Stage II (Phase III), volunteers were required to be >18 years old, healthy or with stable and controlled chronic comorbidities not associated with a reduced immune response according to the investigators' criteria All volunteers were required to have received a complete SARS-CoV-2 vaccination schedule with no more than 3 subsequent booster doses.

##### *Exclusion criteria*

We excluded volunteers with SARS-CoV-2 infection or disease caused by SARS-CoV-2 infection within 90 days before study inclusion, those who had received any vaccine based on an attenuated and inactivated virus or subunits within 28 and 14 days before study inclusion, respectively, those without a complete vaccination regimen against SARS-CoV-2 virus or with more than 3 boosters doses, and those who received the last dose of the primary vaccination regimen or the latest booster within 4 months before study initiation. Volunteers with a scheduled plan to receive any other commercial vaccine within 3 months after study initiation and those who participated in a research study within 60 days before study initiation were also excluded. Volunteers who received an investigational drug or used an invasive investigational medical device in the 30 days prior or received immunoglobulin or investigational monoclonal antibodies in the 3 months before the study's initiation were excluded.

Additional exclusion criteria were a body mass index (BMI) ≥35 kg/m² and medical history of the following conditions: known allergies, anaphylaxis, or any other serious adverse reaction to vaccines or their excipients, clinical condition affecting the function of the immune system, alcoholism or substance abuse, acute polyneuropathy, and any severe or unstable psychiatric condition that would hinder compliance with the protocol's requirements. Volunteers with acute infectious disease at the time of enrollment (excluding minor conditions such as diarrhea or mild upper respiratory tract infection) or a temperature ≥38.0°C in the 24 hours preceding the scheduled vaccination of the study were excluded; admission later was allowed at the investigator's discretion and after consulting with the Sponsor. Volunteers who underwent a surgical procedure that required hospitalization (> 24 hours) in the 12 weeks before vaccination, or those who did not fully recovered from surgery that required hospitalization, or those who had a surgery requiring hospitalization during the expected study participation time or within 6 months after the administration of the investigational vaccine were excluded. We excluded volunteers with any contraindication to receiving intramuscular injections and/or blood extractions, female volunteers who were pregnant or planning to be pregnant within 3 months after the administration of the vaccine, and those in the postpartum period or lactating. Volunteers reporting any condition or finding that, in the investigator's judgment, could jeopardize the subject under investigation or interfere with interpreting the study results were excluded.

##### *Participating Centers*

The eleven participating centers in Argentina were: Centro de Educación Médica e Investigaciones Clínicas (CEMIC), FP Clinical Pharma, Consultorio Médico Itcovici, Vacunar S.A., Fundación Huésped, and Maffei Centro Médico in Buenos Aires, Instituto Médico Platense in La Plata, Centro Clínica del Niño y la Madre and Instituto de Investigaciones Clínicas Mar del Plata in Mar del Plata, ICSAL in Salta, and Clínica Privada del Sol S.A. in Córdoba.

##### *Ethics Committees*

1- Ethics Committee on Clinical Research (CEIC) of the Infectious Studies Center S.A. *Comité de Ética en Investigación Clínica (CEIC) del Centro de Estudios Infectológicos S.A. - CEI – Stamboulian*.

2- Ethics Committee of the Medical Education and Clinical Investigations Center. *Comité de Ética en Investigación de CEMIC.*

3- Research Ethics Committee – Clinical Research Institute Dr. Jesús Vázquez – DiMe Foundation. *Comité de Ética en Investigación - Instituto de Investigaciones Clínicas Dr. Jesús Vázquez - Fundación DiMe.*

4- Provincial Biomedical Research Commission. *Comisión Provincial de Investigaciones Biomédicas.*

5- Research Ethics Committee of the Platense Medical Institute. *Comité de Ética en Investigación Instituto Médico Platense (CEDIMP)*.

6- Institutional Medical Research Ethics Committee -Fabiola- Queen Fabiola University Clinic. *Comité Institucional de Ética en Investigación en Salud CIEIS – Fabiola – Clínica Universitaria Reina Fabiola.*

7- Bioethics Committee (Research Ethics Committee) of the Huésped Foundation. *Comité de Bioética de la Fundación Huésped.*

#### *Randomization Scheme*

For Phase II, we generated the randomization scheme using an online system, available at http://www.randomization.com. We established a system in four blocks of 58 volunteers each, for a total of 232 participants. For Phase III a total of 1782 individuals (1104 aged 18 to 60 years and 678 over 60 years of age), were randomly assigned using a specific software to one of the following three groups: $ARVAC_{Gamma}$ (n=592), $ARVAC_{Omicron}$ (n=594), $ARVAC_{Bivalent}$ (n=594). Each group included 2 subgroups, participants were randomly assigned to receive vaccine first and placebo 28 days later or placebo first and vaccine 28 days later. For the age groups, participants were allocated consecutively until completing the maximum number for each subgroup (184. participants aged 18 to 60 and 113 participants aged >60).

##### *Immunogenicity subset*

The immunogenicity subset included volunteers who have received a complete primary vaccination schedule against the SARS-CoV-2 virus with up to 2 booster doses. Participants with 3 booster doses were not allocated to the immunogenicity subset. Participants with 2 previous booster doses were included in the immunogenicity subset when the time from last booster application was >9 months prior to entry into the study. All participants of the Phase II were allocated to the immunogenicity subset. Whereas in the Phase III, among each subgroup group some participants were randomized to be part of the immunogenicity. In each subgroup that received a vaccine first, 276 participants were allocated to the immunogenicity subset (163 aged 18 to 60 years and 113 from over 60 years) and in each subgroup that received placebo first, 94 participants were allocated to the immunogenicity subset (86 aged 18 to 60 years and 8 over 60 years).

*Secondary Immunogenicity Endpoints*

- Percentage of participants with a ≥8-fold increase from baseline to day 14 in antibody titers to the three SARS-CoV-2 variants.
- Geometric mean of total antibody and neutralizing antibody titers against Ancestral, gamma, and omicron variants and their confidence intervals before and after 14 days of administering a vaccine candidate or placebo, as well as after 90 days of administering a vaccine candidate.
- Comparison of neutralizing antibody levels based on previous SARS-CoV-2 infection history, vaccination platform received in the primary schedule, and age.
- Increase in neutralizing antibody titers compared to baseline against evaluated SARS-CoV-2 variants at 14 days after vaccine or placebo dose and 90 days after vaccine dose.
- Seroconversion rate against different SARS-CoV-2 variants. The seroconversion rate will be assessed as the percentage of subjects who seroconverted after 14 days of receiving the vaccine candidate dose.

Additional exploratory endpoints were levels of total antigen-specific antibodies in blood and saliva at baseline and at 14 days in a selected group.

*NAbs Evaluation*

NAbs against Ancestral, Gamma, and Omicron BA.5 variants were analyzed in the immunogenicity subset of participants who received at least one dose of vaccine or placebo using samples for determination of total and nAbs before (d1) and after the first administration (d14) within an appropriate time frame, and according to the sample size calculation. NAbs against XBB.1.18 and JN.1 SARS-CoV-2 variants were analyzed in samples randomly selected from 87 study participants (48 aged 18-60 and 39 aged >60).

Neutralizing antibodies were evaluated using the reduction of the cytopathic SARS-CoV-2 effect assay in VERO E6 cells (ATCC) as described.[1] Work with SARS-CoV-2 was performed in Biosafety Level 3 laboratories with negative pressure. The ancestral SARS-CoV-2 reference strain 2019 B.1 (GISAID Accession ID: EPI_ISL_499083) was obtained from Sandra Gallegos (Inviv working group); the Gamma SARS-CoV-2 variant (GISAID Accession ID: EPI_ISL_2756556) was isolated by Facundo Di Diego García, Ezequiel Ramírez and Marina Blanco from nasopharyngeal swabs obtained at INBIRS; the Omicron BA.5 (GISAID Accession ID: EPI_ISL_16297058), the XBB.1.18 (GISAID ID EPI_ISL_18746184)and the JN.1 (GISAID Accession ID: EPI_ISL_19037293) SARS-CoV-2 variants were isolated at our laboratory in the University of San Martin from nasopharyngeal swab samples. XBB.1.18 containing nasopharyngeal swab samples were kindly provided by Dario Alvarez from Centro Rossi (Buenos Aires city, Argentina). JN.1 containing nasopharyngeal swab samples were kindly provided by Eugenia Ibañez from Hospital Aleman (Buenos Aires city, Argentina) and Dr. Viegas (Public Health Laboratory, National University of La Plata, Buenos Aires, Argentina). Plasma samples were heat-inactivated (56°C, 30 min) and serial dilutions were incubated with each SARS-CoV-2 variant (1 h, 37°C). The mixture was cultured with VERO E6 cell monolayers for 1 h at 37°C (multiplicity of infection, MOI = 0.01) and replaced by fresh medium. After 72 h, cells were fixed and stained with crystal violet solution in methanol. The virus cytopathic effect on the cell monolayer was then analyzed and the neutralization titer was defined as the highest serum dilution capable of preventing any cytopathic effect.

To analyze exploratory endpoints, anti-Spike and anti-Nucleoprotein IgG-specific antibodies were determined in plasma samples from days 1, 14, and 90, and anti-S1 IgA-specific antibodies were determined in saliva samples from days 1 and 14 using Enzyme-Linked Immunosorbent Assays (ELISA). Anti-Spike IgG SARS-CoV-2 was analyzed using a commercial enzyme-linked immunosorbent microplate assay (COVIDAR – Laboratorio Lemos SRL, Buenos Aires, Argentina), which was calibrated with the World Health Organization (WHO) International Standard, allowing the quantitation in international units (BAU/mL). Nucleoprotein (N)-specific IgG titers were evaluated by indirect ELISA as described in Pasquevich et. al.[1] The anti-SARS-CoV-2 spike S1 IgA levels in saliva were assayed using Anti-SARS-CoV-2 ELISA IgA (Euroimmun, Lübeck, Germany).

##### *Safety Endpoints and Assessments*

Local and systemic adverse events (AEs) were classified according to severity into mild (Grade 1), moderate (Grade 2), severe (Grade 3), and potentially fatal (Grade 4) and according to their relationship to the study medication (related and non-related) based on published guidelines [2]. We also evaluated changes from baseline in basic laboratory parameters 28 (V3) and 56 days (V4) after the first dose administration.

*Sample Size Calculations*

We calculated the sample size with a statistical power >80% (β=0.2), an α=0.05 error, and a 10% dropout rate.

In Phase II, the calculation assumed 85% and 40% seroconversion rates after vaccine and placebo administration, respectively, based on the Phase I study,[1] and was based on two principles: 1) the seroconversion rate at 14 days would be superior after vaccine administration than after placebo administration, 2) it would not be inferior to a 75% reference rate. For the first principle, a sample size of 17 participants in each parallel study arm were deemed necessary, and, for the second principle, the resulting study population was 113 for each vaccine candidate.

In Phase III, the sample size for the immunogenicity analyses was expanded to enable non inferiority and superiority analyses of the seroconversion rates achieved by the bivalent vaccine vs. the monovalent vaccines. For the non-inferiority analysis, the estimated sample size was 158 participants (176 considering a 10% dropout rate) for each vaccine assuming 85% seroconversion rates for both vaccines with a 10% inferiority margin. For the superiority analysis, the estimated sample size was 276 participants for each vaccine, assuming 85% and 75% seroconversion rates for the bivalent and monovalent vaccines, respectively. Based on these calculations, we established a total population of 1342 participants in Phase II and III.

To evaluate the safety endpoints, we based the sample size calculation on two principles: a precision of frequencies not exceeding 3% and detecting at least one AE for those with a 0.1% frequency. For the first principle, we estimated a 50% frequency of AE in our population to ensure an error margin ≤3% regardless of the AEs rates. For the second principle, we considered a statistical power >80% (β=0.2) and an α=0.05 error. According to these premises, 1610 participants (2013 considering a 20% dropout rate) were needed to detect at least one AE with a 0.1% prevalence. To facilitate randomization, we established a safety population of 2014 participants, 232 in Phase I and 1782 in Phase III.

#### *Non-inferiority and Superiority Analyses (Exploratory Endpoints)*

We performed non-inferiority and superiority analyses comparing the $ARVAC_{Bivalent}$ to the $ARVAC_{Gamma}$ and $ARVAC_{Omicron}$ vaccine versions regarding seroconversion rates of nAb titers and the nAb geometric mean titers (GMT) to homologous (i.e., Gamma and Omicron BA.5) and heterologous (Ancestral, Gamma, and Omicron BA.5) SARS-CoV-2 variants.

For the non-inferiority analysis, we defined a <10% non-inferiority margin, whereby for a lower limit of the confidence interval (CI) obtained for the difference between seroconversion rates <10% we would reject the null hypothesis and accept non-inferiority. The 10% margin was transformed into a relative risk (RR) of 0.88, and results were reported as risk differences (RD) and RR, with the 90% CI. The RD, RR and coresponding 90% CIs were calculated using the Wald method.

For the non-inferiority analysis based on GMT ratios (GMTR) we defined a <1.5 inferiority margin in the comparison. $ARVAC_{Gamma}$ and $ARVAC_{Omicron}$ were considered control vaccines and $ARVAC_{Bivalent}$ was considered an experimental vaccine. We calculated the GMTR ($ARVAC_{Gamma}$:$ARVAC_{Bivalent}$ and $ARVAC_{Omicron}$:$ARVAC_{Bivalent}$) and their 90% CI. A situation where the upper 90% CI limit does not cross the 1.5 margin (i.e., <1.5, 50% more nAb in the control vaccines) indicates that the $ARVAC_{Bivalent}$ in not inferior to the $ARVAC_{Gamma}$ or the $ARVAC_{Omicron}$. A situation where the upper 90% CI is >1.5 limit does not allow us to affirm non inferiority. Moreover, an upper limit of the 90% CI <1 may indicate that $ARVAC_{Bivalent}$ is superior. This analysis has a one-tailed type I error of 5%, so the CI may not align with the results of the two-tailed hypothesis tests with a 5% type I error. The results are reported as risk ratios and the 90% CI were estimated using bootstrap (with 2000 replicas). Importantly, the non-inferiority analyses based on GMTR are inverted with respect to the two-tailed superiority analyses. In general, the $ARVAC_{Bivalent}$ induces higher nAb than the other vaccine versions; therefore, we considered that this inversion may facilitate the interpretation of the results, which indicate how many more antibodies participants develop. For non-inferiority analyses, we followed previously described methods.[3]

For the superiority analyses, we compared seroconversion rates between vaccine candidates using the exact Fisher's test and nAbs GMT using the Kruskal Wallis and the Dunns test for multiple comparisons. In addition, we compared nAb GMT ratios (GMTR) between groups using mixed linear regression models. In these models, the dependent variables were the nAb titers of the different SARS-CoV-2 variants at 14 days (on a logarithmic scale), the participant was incorporated as a random term, the vaccine candidates were fixed terms, and the baseline value of nAb titers of each variant was a covariate.

##### *Multivariate Analyses*

Multivariate Poisson regression models for binomial data were used for these analyses, (as described by Voysey *et al.*).[4] The dependent variable for each model was seroconversion for each antibody, and the independent variables were study medication, time since last dose (in days), COVID19 history, age (in years), number of vaccine doses received before the study, and last vaccine platform type. To calculate the 95% CI, the robust variance estimator (sandwich estimator) was used to avoid overestimation of the error with standard methods. The measures of association that emerge from these models (incidence rate ratio) can be interpreted as relative risks adjusted by the variables present in the model. Table 28 shows the distribution of some variables between the groups, as expected for effective randomization, there are no statistically significant differences between the groups.

##### *Additional Statistical Methods*

Categorical variables were described as frequencies and percentages, and quantitative variables were described with the mean and standard deviation (SD) and the median and quartiles 1 and 3 (Q1, Q3; interquartile range, IQR).

Categorical variables were compared using the Fisher's or the Chi-square test. NAb levels were compared between timepoints using the non-parametric two-tailed paired Friedman test and the Dunns test, or the non-parametric paired Wilcoxon test.

The R statistical packages used in the analyses were tidyverse, ggplot2, gtsummary, dani, TOSTER, DescTools, gt, ggpubr, ggsci, rio, pubh, and wesanderson.

2. US Food and Drug Administration. Guidance for industry: toxicity grading scale for healthy adult and adolescent volunteers enrolled in preventive vaccine clinical trials. 2007.

3. Corominas J, Garriga C, Prenafeta A, et al. Safety and immunogenicity of the protein-based PHH-1V compared to BNT162b2 as a heterologous SARS-CoV-2

booster vaccine in adults vaccinated against COVID-19: a multicentre, randomised, double-blind, non-inferiority phase IIb trial. The Lancet regional health Europe [Internet] 2023 [cited 2024 Mar 19];28. Available from: https://pubmed.ncbi.nlm.nih.gov/37131861/

4. Voysey M, Ann S, Clemens C, et al. Safety and efficacy of the ChAdOx1 nCoV-19 vaccine (AZD1222) against SARS-CoV-2: an interim analysis of four randomised controlled trials in Brazil, South Africa, and the UK. www.thelancet.com [Internet] 2021 [cited 2024 Apr 24];397:2021. Available from: https://doi.org/10.1016/

### Supplementary Tables

**Table S1.** Characteristics of Phase III participants included in the immunogenicity analysis. N=1053

| | | | ARVAC | | |
|---|---|---|---|---|---|
| | **Placebo** | **Gamma** | **Omicron BA.4/5** | **Bivalent** | **Total** |
| **Participants included in the analysis, n** | | | | | |
| *All participants* | 264 | 265 | 265 | 259 | 1053 |
| *Participants 18-60 years* | 158 | 156 | 157 | 157 | 628 |
| *Participants >60 years* | 106 | 109 | 108 | 102 | 425 |
| **Age (years),** ***median (IQR)*** | | | | | |
| *All participants* | 53.5 (38.8, 64.0) | 53.0 (36.0, 63.0) | 52.0 (36.0, 65.0) | 50.0 (36.0, 64.0) | 52.0 (36.0, 64.0) |
| *Participants 18-60 years* | 42.5 (31.0, 50.8) | 40.0 (27.0, 48.3) | 40.0 (28.0, 49.0) | 40.0 (28.0, 48.0) | 41.0 (28.0, 49.0) |
| *Participants >60 years* | 66.0 (63.0, 69.0) | 65.0 (62.0, 70.0) | 66.5 (63.0, 70.0) | 65.0 (62.0, 69.0) | 65.0 (63.0, 70.0) |
| **Sex,** ***n (%)*** | | | | | |
| *All participants* | | | | | |
| Women | 115 (43.6) | 121 (45.7) | 125 (47.2) | 126 (48.6) | 487 (46.2) |
| Men | 149 (56.4) | 144 (54.3) | 140 (52.8) | 133 (51.4) | 566 (53.8) |
| *Participants 18-60 years* | | | | | |
| Women | 72 (45.6) | 76 (48.7) | 75 (47.8) | 92 (58.6) | 315 (50.2) |
| Men | 86.0 (54.4) | 80 (51.3) | 82 (52.2) | 65 (41.4) | 313 (49.8) |
| *Participants >60 years* | | | | | |
| Women | 43 (40.6) | 45 (41.3) | 50 (46.3) | 34 (33.3) | 172 (40.5) |
| Men | 63 (59.4) | 64 (58.7) | 58 (53.7) | 68 (66.7) | 253 (59.5) |
| **Body mass index (kg/m$^2$),** ***median (IQR)*** | | | | | |
| *All participants* | 26.8 (23.7, 30.4) | 26.9 (23.9, 29.6) | 27.5 (23.6, 30.4) | 26.9 (23.6, 29.8) | 27.1 (23.7, 30.1) |
| *Participants 18-60 years* | 26.4 (23.3, 30.5) | 26.5 (23.3, 29.2) | 27.2 (23.6, 29.9) | 25.6 (22.5, 28.9) | 26.5 (23.1, 29.4) |
| *Participants >60 years* | 27.7 (24.3, 30.3) | 27.2 (24.7, 30.7) | 28.1 (23.8, 31.0) | 28.4 (24.9, 31.7) | 27.9 (24.6, 31.0) |
| **Participants with comorbidities,** ***n (%)*** | | | | | |
| All participants | 134.0 (50.8) | 134.0 (50.6) | 136.0 (51.3) | 144.0 (55.6) | 548.0 (52.0) |
| Participants 18-60 years | 58.0 (36.7) | 61.0 (39.51) | 65.0 (41.4) | 64.0 (40.8) | 249.0 (39.6) |
| Participants >60 years | 76.0 (71.7) | 73.0 (67.0) | 71.0 (65.7) | 80.0 (78.4) | 299.0 (70.4) |
| **Time since last vaccine administration (days),** ***median (IQR)*** | | | | | |
| All participants | 509.5 (455.0, 580.0) | 503.0 (455.0, 569.0) | 504.0 (454, 558) | 497 (455.0, 549.5) | 504.0 (454.0, 565.0) |
| Participants 18-60 years | 471.0 (436.3, 512.8) | 475.5 (427.0, 505.3) | 474.5 (423, 514) | 475 (436.0, 515.0) | 474.0 (430.0, 511.3) |
| Participants >60 years | 564.0 (529.0, 594.0) | 552.0 (514.0, 593.0) | 540.0 (514.5, 588) | 543 (497.5, 580.8) | 549.0 (515.0, 592.0) |
| **Number of booster doses,** ***n (%)*** | | | | | |
| ***All participants*** | 264 (100.0) | 265 (100.0) | 265 (100.0) | 259 (100.0) | 1053 (100.0) |
| No booster | 34 (12.9) | 32 (12.1) | 38 (14.3) | 33 (12.7) | 137 (13.0) |
| One booster | 183 (69.3) | 189 (71.3) | 188 (70.9) | 194 (74.9) | 754 (71.6) |
| Two boosters | 47 (17.8) | 44 (16.6) | 39 (14.7) | 32 (12.4) | 162 (15.4) |

| | | | | | |
|---|---|---|---|---|---|
| ***Participants 18-60 years*** | 158 (100.0) | 156 (100.0) | 156 (100.0) | 157 (100.0) | 628 (100.0) |
| No booster | 27 (17.1) | 23 (14.7) | 30 (19.1) | 27 (17.2) | 107 (17.0) |
| One booster | 131 (82.9) | 131 (84.0) | 126 (80.3) | 130 (82.8) | 518 (82.5) |
| Two boosters | 0 (0.0) | 2 (1.3) | 1 (0.6) | 0 (0.0) | 3 (0.5) |
| ***Participants >60 years*** | 106 (100.0) | 109 (100.0) | 108 (100.0) | 102 (100.0) | 425 (100.0) |
| No booster | 7 (6.6) | 9 (8.3) | 8 (7.4) | 6 (5.9) | 30 (7.1) |
| One booster | 52 (49.1) | 58 (53.2) | 62 (57.4) | 64 (62.7) | 236 (55.5) |
| Two boosters | 47 (44.3) | 42 (38.5) | 38 (35.2) | 32 (31.4) | 159 (37.4) |
| **Primary vaccine platform, *n (%)*** | | | | | |
| ***All participants*** | | | | | |
| Adenovirus | 150 (56.8) | 134 (50.8) | 151 (57.0) | 143 (55.2) | 578 (54.9) |
| Adenovirus monodosis | 13 (4.9) | 14 (5.3) | 9 (3.4) | 6 (2.3) | 42 (4.0) |
| mRNA | 7 (2.7) | 7 (2.7) | 12 (4.5) | 13 (5.0) | 39 (3.7) |
| Inactivated vaccine | 56 (21.2) | 62 (23.5) | 46 (17.4) | 55 (21.2) | 219 (20.8) |
| Heterologous platforms | 36 (13.6) | 45 (17.0) | 47 (17.7) | 42 (16.2) | 170 (16.2) |
| Recombinant protein | 2 (0.8) | 2 (0.8) | 0 (0.0) | 0 (0.0) | 4 (0.4) |
| ***Participants 18-60 years*** | | | | | |
| Adenovirus | 84 (53.2) | 71 (45.8) | 78 (49.7) | 78 (49.7) | 311 (49.5) |
| Adenovirus monodosis | 8 (5.1) | 10 (6.5) | 8 (5.1) | 6 (3.8) | 32 (5.1) |
| mRNA | 6 (3.8) | 7 (4.5) | 8 (5.1) | 11 (7.0) | 32 (5.1) |
| Inactivated vaccine | 48 (30.4) | 50 (32.3) | 41 (26.1) | 48 (30.6) | 188 (29.9) |
| Heterologous platforms | 11 (7.0) | 15 (9.7) | 22 (14.0) | 14 (8.9) | 62 (9.9) |
| Recombinant protein | 1 (0.6) | 2 (1.3) | 0 (0.0) | 0 (0.0) | 3 (0.5) |
| ***Participants >60 years*** | | | | | |
| Adenovirus | 66 (62.3) | 63 (57.8) | 73 (67.6) | 65 (63.7) | 267 (62.8) |
| Adenovirus monodosis | 5 (4.7) | 4 (3.7) | 1 (0.9) | 0 (0.0) | 10 (2.4) |
| mRNA | 1 (0.9) | 0 (0.0) | 4 (3.7) | 2 (2.0) | 7 (1.6) |
| Inactivated vaccine | 8 (7.5) | 12 (11.0) | 5 (4.6) | 7 (6.9) | 32 (7.5) |
| Heterologous platforms | 25 (23.6) | 30 (27.5) | 25 (23.1) | 28 (27.5) | 108 (25.4) |
| Recombinant protein | 1 (0.9) | 0 (0.0) | 0 (0.0) | 0 (0.0) | 1 (0.2) |

IQR, interquartile range

**Table S2.** Seroconversion rates using normalized antibody titers in Phase III participants for the different vaccine variants and age groups compared to placebo and a >75% reference. n=1053

| Study Phase and treatment | SARS-CoV-2 variant | Seroconversion rate (%) | 95% CI | P-value[a] vaccine vs. placebo | P-value[b] vaccine vs. >75% |
|---|---|---|---|---|---|
| ***All participants, n=1053*** | | | | | |
| Placebo, n= 264 | Ancestral | 12.5 | 9.0-17.0 | NA | NA |
| | Gamma | 12.1 | 8.7-16.6 | NA | NA |
| | Omicron BA.5 | 15.2 | 11.3-20.0 | NA | NA |
| ARVAC Gamma, n=265 | Ancestral | 86.8 | 82.2-90.3 | <.0001 | <.0001 |
| | Gamma | 84.2 | 79.3-88.1 | <.0001 | .0006 |
| | Omicron BA.5 | 81.9 | 76.8-86.1 | <.0001 | .010 |
| ARVAC Omicron, n=265 | Ancestral | 80.0 | 74.8-84.4 | <.0001 | .06 |
| | Gamma | 82.3 | 77.2-86.4 | <.0001 | .006 |
| | Omicron BA.5 | 87.5 | 83.0-91.0 | <.0001 | <.0001 |
| ARVAC Bivalent, n=259 | Ancestral | 92.3 | 88.4-94.9 | <.0001 | <.0001 |
| | Gamma | 91.1 | 87.0-94.0 | <.0001 | <.0001 |
| | Omicron BA.5 | 92.7 | 88.8-95.3 | <.0001 | <.0001 |
| ***Participants 18-60 years*** | | | | | |
| Placebo, n= 157 | Ancestral | 7.6 | 4.4-12.9 | NA | NA |
| | Gamma | 9.6 | 5.9-15.2 | NA | NA |
| | Omicron BA.5 | 14.0 | 9.4-20.3 | NA | NA |
| ARVAC Gamma, n=156 | Ancestral | 89.1 | 83.2-93.1 | <.0001 | <.0001 |
| | Gamma | 89.1 | 83.2-93.1 | <.0001 | <.0001 |
| | Omicron BA.5 | 86.5 | 80.3-91.0 | <.0001 | .0009 |
| ARVAC Omicron, n=156 | Ancestral | 81.5 | 74.7-86.8 | <.0001 | .06 |
| | Gamma | 86.6 | 80.4-91.1 | <.0001 | .0008 |
| | Omicron BA.5 | 87.3 | 81.1-91.6 | <.0001 | .0004 |
| ARVAC Bivalent, n=156 | Ancestral | 93.0 | 87.9-96.0 | <.0001 | <.0001 |
| | Gamma | 91.7 | 86.3-95.1 | <.0001 | <.0001 |
| | Omicron BA.5 | 92.4 | 87.1-95.6 | <.0001 | <.0001 |
| ***Participants >60 years*** | | | | | |
| Placebo, n= 106 | Ancestral | 19.8 | 13.3-28.4 | NA | NA |
| | Gamma | 16.0 | 10.3-24.2 | NA | NA |
| | Omicron BA.5 | 17.9 | 11.8-26.3 | NA | NA |
| ARVAC Gamma, n=109 | Ancestral | 83.5 | 75.4-89.3 | <.0001 | .04 |
| | Gamma | 77.1 | 68.3-84.0 | <.0001 | .62 |
| | Omicron BA.5 | 75.2 | 66.4-82.4 | <.0001 | .96 |
| ARVAC Omicron, n=108 | Ancestral | 77.8 | 69.1-84.6 | <.0001 | .504 |
| | Gamma | 75.9 | 67.1-83.0 | <.0001 | .82 |
| | Omicron BA.5 | 88.0 | 80.5-92.8 | <.0001 | .002 |
| ARVAC Bivalent, n=102 | Ancestral | 90.2 | 82.9-94.6 | <.0001 | .0004 |
| | Gamma | 90.2 | 82.9-94.6 | <.0001 | .0004 |
| | Omicron BA.5 | 92.2 | 85.3-96.0 | <.0001 | <.0001 |

CI, confidence interval; NA, not applicable.
[a]Calculated using the Fisher's exact test.
[b]Calculated using the Z-test.

**Table S3.** Percentages of participants in Phase II (N=228) and Phase III (n=1053) with ≥8-fold increases in neutralizing antibody titers after vaccine or placebo administration.

| Treatment | SARS-CoV-2 variant | % | 95% CI | P-value vaccine vs. placebo[a] |
|---|---|---|---|---|
| **Phase II, n=228** | | | | |
| Placebo, n=114 | Ancestral | 3.5 | 1.4-8.7 | NA |
| | Gamma | 3.5 | 1.4-8.7 | NA |
| | Omicron BA.5 | 1.8 | 0.3-6.2 | NA |
| ARVAC Gamma, n=114 | Ancestral | 56.1 | 47.0-64.9 | <.0001 |
| | Gamma | 62.3 | 53.1-70.6 | <.0001 |
| | Omicron BA.5 | 55.3 | 46.1-64.1 | <.0001 |
| **Phase III all participants, n=1053** | | | | |
| Placebo, n= 264 | Ancestral | 1.5 | 0.6-3.8 | NA |
| | Gamma | 1.5 | 0.6-3.8 | NA |
| | Omicron BA.5 | 2.7 | 1.3-5.4 | NA |
| ARVAC Gamma, n=265 | Ancestral | 61.1 | 55.1-66.8 | <.0001 |
| | Gamma | 63.4 | 57.4-69.0 | <.0001 |
| | Omicron BA.5 | 56.6 | 50.6-62.4 | <.0001 |
| ARVAC Omicron, n=265 | Ancestral | 53.2 | 47.2-59.1 | <.0001 |
| | Gamma | 54.7 | 48.7-60.6 | <.0001 |
| | Omicron BA.5 | 66.8 | 60.9-72.2 | <.0001 |
| ARVAC Bivalent, n=259 | Ancestral | 70.7 | 64.8-75.9 | <.0001 |
| | Gamma | 69.5 | 63.6-74.8 | <.0001 |
| | Omicron BA.5 | 75.3 | 69.7-80.1 | <.0001 |

CI, confidence interval.
[a]Calculated using the Fisher's exact test.

**Table S4.** Non-inferiority analysis for seroconversion rates of ARVAC Bivalent vs. ARVAC Gamma and ARVAC Omicron in all Phase III participants.

| Comparisons | Risk difference | 90% CI[a] | Non-inferiority p-value[b] | Relative risk | 90% CI[a] | Non-inferiority p-valueb |
|---|---|---|---|---|---|---|
| ***All Phase III participants*** | | | | | | |
| **Ancestral variant** | | | | | | |
| Bivalent vs. Omicron | 12.66 | 7.44-17.89 | <.001 | 1.16 | 1.10-1.24 | <.001 |
| Bivalent vs. Gamma | 5.87 | 1.15-10.59 | <.001 | 1.07 | 1.01-1.13 | <.001 |
| **Gamma variant** | | | | | | |
| Bivalent vs. Omicron | 8.85 | 3.64-14.07 | <.001 | 1.11 | 1.04-1.18 | <.001 |
| Bivalent vs. Gamma | 6.97 | 1.89-12.05 | <.001 | 1.08 | 1.02-1.15 | <.001 |
| **Omicron BA.5 variant** | | | | | | |
| Bivalent vs. Omicron | 5.12 | 0.47-9.77 | <.001 | 1.06 | 1.01-1.12 | <.001 |
| Bivalent vs. Gamma | 10.78 | 5.68-15.87 | <.001 | 1.13 | 1.07-1.20 | <.001 |
| ***Participants 18-60 years*** | | | | | | |
| **Ancestral variant** | | | | | | |
| Bivalent vs. Omicron | 12.1 | 5.45-18.76 | <.001 | 1.15 | 1.06-1.25 | <.001 |
| Bivlent vs. Gamma | 4.53 | -1.32-10.37 | <.001 | 1.05 | 0.98-1.13 | <.001 |
| **Gamma variant** | | | | | | |
| Bivalent vs. Omicron | 4.46 | -1.86-10.77 | <.001 | 1.05 | 0.98-1.13 | <.001 |
| Bivalent vs. Gamma | 2.62 | -3.49-8.73 | <.001 | 1.03 | 0.96-1.10 | <.001 |
| **Omicron BA.5 variant** | | | | | | |
| Bivalent vs. Omicron | 5.73 | -0.04-11.88 | <.001 | 1.07 | 0.99-1.15 | <.001 |
| Bivalent vs. Gamma | 6.45 | 0.21-12.70 | <.001 | 1.07 | 1.00-1.16 | <.001 |
| ***Participants >60 years*** | | | | | | |
| **Ancestral variant** | | | | | | |
| Bivalent vs. Omicron | 13.4 | 4.41-22.39 | <.001 | 1.17 | 1.05-1.32 | <.001 |
| Bivalent vs. Gamma | 7.69 | -0.71-16.09 | <.001 | 1.09 | 0.99-1.21 | <.001 |
| **Gamma variant** | | | | | | |
| Bivalent vs. Omicron | 15.2 | 5.85-24.54 | <.001 | 1.2 | 1.07-1.36 | <.001 |
| Bivalent vs. Gamma | 13.13 | 3.98-22.29 | <.001 | 1.17 | 1.05-1.32 | <.001 |
| **Omicron BA.5 variant** | | | | | | |
| Bivalent vs. Omicron | 4.19 | -0.03-11.91 | <.001 | 1.07 | 0.99-1.15 | <.001 |
| Bivalent vs. Gamma | 16.93 | 7.89-25.96 | <.001 | 1.07 | 1.00-1.16 | <.001 |

CI, confidence interval

[a]Estimated using the Wald method

[b]Calculated using the Wald test with one-tailed continuity correction with a 5% type I error

**Table S5.** Superiority comparisons between vaccine versions in Phase III participants.

| SARS-CoV-2 variant | Seroconversion rates (%) | | | Adjusted p-values[a] | | |
|---|---|---|---|---|---|---|
| | Bivalent | Gamma | Omicron | Bivalent vs. Gamma | Bvialent vs. Omicron | Gamma vs. Omicron |
| ***All participants*** | *n=259* | *n=265* | *n=265* | | | |
| Ancestral | 92.7 | 86.8 | 80.0 | .116 | 0 | .142 |
| Omicron BA.5 | 92.7 | 81.9 | 87.5 | .001 | .21 | .273 |
| Gamma | 91.1 | 84.2 | 82.3 | .067 | .013 | 1 |
| ***Participants 18-60 years*** | *n=157* | *n=156* | *n=157* | | | |
| Ancestral | 93.6 | 89.1 | 81.5 | .661 | .006 | .25 |
| Omicron BA.5 | 93.0 | 86.5 | 87.3 | .268 | .39 | 1 |
| Gamma | 91.7 | 89.1 | 87.3 | 1 | .809 | 1 |
| ***Participants >60 years*** | *n=102* | *n=109* | *n=108* | | | |
| Ancestral | 91.2 | 83.5 | 77.8 | .429 | .04 | 1 |
| Omicron BA.5 | 92.2 | 75.2 | 88.0 | .005 | 1 | .075 |
| Gamma | 90.2 | 77.1 | 75.0 | .052 | .02 | 1 |

[a]Adjusted using the Bonferroni method.

**Table S6.** Multivariate models comparing seroconversion to the Ancestral variant between vaccine versions.

| | Bivalent vs. Omicron | | | Bivalent vs. Gamma | | | Gamma vs. Omicron | | |
|---|---|---|---|---|---|---|---|---|---|
| **Co-variates** | **Incidence rate ratio** | **95% CI** | **P-value** | **Incidence rate ratio** | **95% CI** | **P-value** | **Incidence rate ratio** | **95% CI** | **P-value** |
| **Vaccine** | | | | | | | | | |
| Omicron | — | — | | | | | 0.92 | 0.86-1.00 | .044 |
| Bivalent | 1.16 | 1.08-1.24 | <.001 | 1.07 | 1.01-1.13 | .03 | | | |
| Gamma | | | | — | — | | — | — | |
| **Days since last dose** | 1 | 1.00-1.00 | .042 | 1 | 1.00-1.00 | .7 | 1 | 1.00-1.00 | .2 |
| **Age** | 1 | 1.00-1.00 | .028 | 1 | 1.00-1.00 | .2 | 1 | 0.99-1.00 | .075 |
| **Previous COVID-19** | | | | | | | | | |
| No | — | — | | — | — | | — | — | |
| Yes | 1.02 | 0.95-1.09 | .6 | 0.98 | 0.92-1.05 | .6 | 1 | 0.92-1.07 | .9 |
| **Number of previous vaccines** | 1.01 | 0.94-1.09 | .7 | 1.01 | 0.94-1.08 | .8 | 1.04 | 0.95-1.13 | .4 |
| **Vaccine platform - last dose** | | | | | | | | | |
| mRNA | — | — | | — | — | | — | — | |
| Viral vector | 0.98 | 0.91-1.06 | .7 | 0.98 | 0.91-1.06 | .6 | 1.02 | 0.93-1.11 | .6 |
| Inactivated virus | 1.01 | 0.87-1.17 | .9 | 1.08 | 0.97-1.21 | .2 | 1.08 | 0.91-1.28 | .4 |
| Protein | | | | 1.14 | 1.00-1.29 | .052 | 1.14 | 0.99-1.32 | .065 |

CI, confidence interval

**Table S7**. Multivariate models comparing seroconversion to the Gamma variant between vaccine versions.

| Co-variates | Bivalent vs. Omicron | | | Bivalent vs. Gamma | | | Gamma vs. Omicron | | |
|---|---|---|---|---|---|---|---|---|---|
| | Incidence rate ratio | 95% CI | P-value | Incidence rate ratio | 95% CI | P-value | Incidence rate ratio | 95% CI | P-value |
| **Vaccine** | | | | | | | | | |
| Omicron | — | — | | | | | 0.99 | 0.91-1.06 | .7 |
| Bivalent | 1.1 | 1.03-1.17 | .005 | 1.08 | 1.02-1.15 | .015 | | | |
| Gamma | | | | — | — | | — | — | |
| **Days since last dose** | 1 | 1.00-1.00 | .015 | 1 | 1.00-1.00 | .004 | 1 | 1.00-1.00 | .009 |
| **Age** | 1 | 0.99-1.00 | <.001 | 1 | 0.99-1.00 | .004 | 0.99 | 0.99-1.00 | <.001 |
| **Previous COVID-19** | | | | | | | | | |
| No | — | — | | — | — | | — | — | |
| Yes | 0.99 | 0.92-1.05 | .7 | 0.97 | 0.91-1.03 | .3 | 1 | 0.92-1.07 | >.9 |
| **Number of previous vaccines** | 1 | 0.93-1.09 | >.9 | 1 | 0.94-1.06 | >.9 | 1.02 | 0.94-1.09 | .7 |
| **Vaccine platform - last dose** | | | | | | | | | |
| mRNA | — | — | | — | — | | — | — | |
| Viral vector | 0.96 | 0.89-1.03 | .3 | 0.99 | 0.92-1.06 | .8 | 0.98 | 0.90-1.07 | .7 |
| Inactivated virus | 0.61 | 0.42-0.88 | .008 | 0.86 | 0.69-1.07 | .2 | 0.7 | 0.51-0.97 | .03 |
| Protein | | | | 1.06 | 0.95-1.17 | .3 | 1.08 | 0.95-1.22 | .2 |

CI, confidence interval

**Table S8.** Multivariate models comparing seroconversion to the Omicron variant between vaccine versions.

| | Bivalent vs. Omicron | | | Bivalent vs. Gamma | | | Gamma vs. Omicron | | |
|---|---|---|---|---|---|---|---|---|---|
| **Co-variates** | **Incidence rate ratio** | **95% CI** | **P-value** | **Incidence rate ratio** | **95% CI** | **P-value** | **Incidence rate ratio** | **95% CI** | **P-value** |
| **Vaccine** | | | | | | | | | |
| Omicron | — | — | | | | | 1.07 | 1.00-1.15 | .054 |
| Bivalent | 1.06 | 1.00-1.12 | .054 | 1.13 | 1.06-1.21 | <.001 | | | |
| Gamma | | | | — | — | | — | — | |
| **Days since last dose** | 1 | 1.00-1.00 | .028 | 1 | 1.00-1.00 | .4 | 1 | 1.00-1.00 | .2 |
| **Age** | 1 | 1.00-1.00 | .071 | 1 | 0.99-1.00 | .008 | 1 | 0.99-1.00 | .043 |
| **Previous COVID-19** | | | | | | | | | |
| No | — | — | | — | — | | — | — | |
| Yes | 1.01 | 0.95-1.06 | .8 | 0.99 | 0.92-1.06 | .7 | 0.98 | 0.91-1.06 | .6 |
| **Number of previous vaccines** | 1.03 | 0.97-1.10 | .3 | 1.02 | 0.94-1.11 | .6 | 1.01 | 0.93-1.10 | .8 |
| **Vaccine platform - last dose** | | | | | | | | | |
| mRNA | — | — | | — | — | | — | — | |
| Viral vector | 0.95 | 0.88-1.02 | .2 | 0.99 | 0.92-1.07 | .8 | 1.02 | 0.94-1.11 | .7 |
| Inactivated virus | 0.93 | 0.77-1.11 | .4 | 1 | 0.82-1.21 | >.9 | 1.02 | 0.86-1.22 | .8 |
| Protein | | | | 1.2 | 1.03-1.39 | .02 | 1.17 | 1.01-1.35 | .036 |

CI, confidence interval

**Table S9.** Non-inferiority analysis of geometric mean titers ratios (GMTR).

| Comparisons | GMTR | 90% CI | Non-inferiority *p*-value |
|---|---|---|---|
| ***All participants*** | | | |
| **Ancestral** | | | |
| Bivalent vs Omicron | 0.83 | 0.68-1.01 | <.001 |
| Bivalent vs Gamma | 0.84 | 0.68-1.04 | <.001 |
| **Gamma** | | | |
| Bivalent vs Omicron | 0.79 | 0.64-0.96 | <.001 |
| Bivalent vs Gamma | 0.84 | 0.70-1.04 | <.001 |
| **Omicron BA.5** | | | |
| Bivalent vs Omicron | 0.93 | 0.75-1.15 | <.001 |
| Bivalent vs Gamma | 0.54 | 0.44-0.67 | <.001 |
| ***Participants 18-60 years*** | | | |
| **Ancestral** | | | |
| Bivalent vs Omicron | 0.81 | 0.62-1.06 | <.001 |
| Bivalent vs Gamma | 0.9 | 0.69-1.15 | <.001 |
| **Gamma** | | | |
| Bivalent vs Omicron | 0.87 | 0.67-1.11 | <.001 |
| Bivalent vs Gamma | 1.01 | 0.78-1.27 | .003 |
| **Omicron BA.5** | | | |
| Bivalent vs Omicron | 0.93 | 0.69-1.25 | .003 |
| Bivalent vs Gamma | 0.63 | 0.49-0.81 | <.001 |
| ***Participants >60 years*** | | | |
| **Ancestral** | | | |
| Bivalent vs Omicron | 0.85 | 0.62-1.13 | <.001 |
| Bivalent vs Gamma | 0.74 | 0.54-1.04 | <.001 |
| **Gamma** | | | |
| Bivalent vs Omicron | 0.68 | 0.49-0.92 | <.001 |
| Bivalent vs Gamma | 0.65 | 0.45-0.92 | <.001 |
| **Omicron BA.5** | | | |
| Bivalent vs Omicron | 0.93 | 0.67-1.27 | .006 |
| Bivalent vs Gamma | 0.43 | 0.30-0.61 | <.001 |

CI, confidence interval; GMTR, geometric mean titers ratios.

**Table S10.** Comparison of geometric mean titers (GMT) between groups before and after placebo or vaccine administration in all Phase III participants. N=1053.

| SARS-CoV-2 variant and day | Placebo (N=264) GMT (95% CI) | Gamma (N=265) GMT (95% CI) | Omicron (N=265) GMT (95% CI) | Bivalent (N=259) GMT (95% CI) | Comparisons between treatments | Significance | Adjusted *p*-value[a] |
|---|---|---|---|---|---|---|---|
| **Ancestral** | | | | | | | |
| d1 | 125.3 (105.7-148.6) | 127.7 (106.9-152.5) | 162.4 (137.6-191.7) | 121.3 (103.1-142.8) | Placebo d1 vs. ARVAC Bivalent d1 | ns | >.9999 |
| | | | | | Placebo d1 vs. ARVAC Omicron d1 | ns | .2377 |
| | | | | | Placebo d1 vs. ARVAC Gamma d1 | ns | >.9999 |
| | | | | | ARVAC Bivalent d1 vs. ARVAC Omicron d1 | ns | .1121 |
| | | | | | ARVAC Bivalent d1 vs. ARVAC Gamma d1 | ns | >.9999 |
| | | | | | ARVAC Omicron vs. ARVAC Gamma d1 | ns | .2585 |
| d14 | 131.4 (111.2-155.3) | 1227 (1030-1461) | 1214 (1021-1443) | 1462 (1228-1740) | Placebo d14 vs. ARVAC Bivalent d14 | **** | <.0001 |
| | | | | | Placebo d14 vs. ARVAC Omicron d14 | **** | <.0001 |
| | | | | | Placebo d14 vs. ARVAC Gamma d14 | **** | <.0001 |
| | | | | | ARVAC Bivalent d14 vs. ARVAC Omicron d14 | ns | .9339 |
| | | | | | ARVAC Bivalent d14 vs. ARVAC Gamma d14 | ns | .8491 |
| | | | | | Omicron d14 vs. ARVAC Gamma d14 | ns | >.9999 |
| **Gamma** | | | | | | | |
| d1 | 94.15 (79.17-112) | 87.37 (73.52-103.8) | 101.4 (86.24-119.3) | 87.06 (74.22-102.1) | Placebo d1 vs. ARVAC Bivalent d1 | ns | >.9999 |
| | | | | | Placebo d1 vs. ARVAC Omicron d1 | ns | >.9999 |
| | | | | | Placebo d1 vs. ARVAC Gamma d1 | ns | >.9999 |
| | | | | | ARVAC Bivalent d1 vs. ARVAC Omicron d1 | ns | >.9999 |
| | | | | | ARVAC Bivalent d1 vs. ARVAC Gamma d1 | ns | >.9999 |
| | | | | | ARVAC Omicron d1 vs. ARVAC Gamma d1 | ns | >.9999 |
| d14 | 97.67 (82.24-116) | 900.8 (755.9-1074) | 839.4 (706.4-997.5) | 1066 (898.9-1264) | Placebo d14 vs. ARVAC Bivalent d14 | **** | <.0001 |
| | | | | | Placebo d14 vs. ARVAC Omicron d14 | **** | <.0001 |
| | | | | | Placebo d14 vs. ARVAC Gamma d14 | **** | <.0001 |
| | | | | | ARVAC Bivalent d14 vs. ARVAC Omicron d14 | ns | .9339 |
| | | | | | ARVAC Bivalent d14 vs. ARVAC Gamma d14 | ns | .8491 |
| | | | | | ARVAC Omicron d14 vs. ARVAC Gamma d14 | ns | >.9999 |
| **Omicron BA.5** | | | | | | | |
| d1 | 47.57 (40.22-56.26) | 51.11 (42.73-61.13) | 58.86 (49.93-69.38) | 51.53 (43.8-60.61) | Placebo d1 vs. ARVAC Bivalent d1 | ns | >.9999 |
| | | | | | Placebo d1 vs. ARVAC Omicron d1 | ns | .8841 |
| | | | | | Placebo d1 vs. ARVAC Gamma d1 | ns | >.9999 |
| | | | | | ARVAC Bivalent d1 vs. ARVAC Omicron d1 | ns | >.9999 |
| | | | | | ARVAC Bivalent d1 vs. ARVAC Gamma d1 | ns | >.9999 |
| | | | | | ARVAC Omicron d1 vs. ARVAC Gamma d1 | ns | >.9999 |
| d14 | 49.74 (41.85-59.11) | 426.3 (356.4-510.1) | 734.6 (614-878.9) | 789.9 (661-943.9) | Placebo d14 vs. ARVAC Bivalent d14 | **** | <.0001 |
| | | | | | Placebo d14 vs. ARVAC Omicron d14 | **** | <.0001 |
| | | | | | Placebo d14 vs. ARVAC Gamma d14 | **** | <.0001 |
| | | | | | ARVAC Bivalent d14 vs. ARVAC Omicron d14 | ns | >.9999 |
| | | | | | ARVAC Bivalent d14 vs. ARVAC Gamma d14 | *** | .0002 |
| | | | | | ARVAC Omicron d14 vs. ARVAC Gamma d14 | ** | .0033 |

CI, confidence interval; GMT, geometric mean titers; ns, not significant.
[a]Non-parametric two-tailed Kruskal Wallis test for unpaired data and Dunns test for multiple comparisons.

**Table S11.** Comparison of geometric mean titers (GMT) between groups before and after placebo or vaccine administration in Phase III participants aged 18-60 years. N=628.

| SARS-CoV-2 variant and day | Placebo (N=158) GMT (95% CI) | Gamma (N=156) GMT (95% CI) | Omicron (N=157) GMT (95% CI) | Bivalent (N=157) GMT (95% CI) | Comparisons between treatments | Significance | Adjusted *p*-value[a] |
|---|---|---|---|---|---|---|---|
| **Ancestral** | | | | | | | |
| d1 | 101.4 (83.03-123.9) | 114 (91.27-142.5) | 136.8 (110.8-168.8) | 101.3 (83.28-123.2) | Placebo d1 vs. ARVAC Bivalent d1 | ns | >.9999 |
| | | | | | Placebo d1 vs. ARVAC Omicron d1 | ns | .3695 |
| | | | | | Placebo d1 vs. ARVAC Gamma d1 | ns | >.9999 |
| | | | | | ARVAC Bivalent d1 vs. ARVAC Omicron d1 | ns | .2207 |
| | | | | | ARVAC Bivalent d1 vs. ARVAC Gamma d1 | ns | >.9999 |
| | | | | | ARVAC Omicron vs. ARVAC Gamma d1 | ns | .8374 |
| d14 | 101.9 (84.65-122.6) | 1139 (923.5-1405) | 1024 (811.6-1292) | 1260 (1004-1582) | Placebo d14 vs. ARVAC Bivalent d14 | **** | <.0001 |
| | | | | | Placebo d14 vs. ARVAC Omicron d14 | **** | <.0001 |
| | | | | | Placebo d14 vs. ARVAC Gamma d14 | **** | <.0001 |
| | | | | | ARVAC Bivalent d14 vs. ARVAC Omicron d14 | ns | >.9999 |
| | | | | | ARVAC Bivalent d14 vs. ARVAC Gamma d14 | ns | >.9999 |
| | | | | | ARVAC Omicron d14 vs. ARVAC Gamma d14 | ns | >.9999 |
| **Gamma** | | | | | | | |
| d1 | 76.28 (62.4-93.24) | 76.45 (61.42-95.15) | 91.51 (74.66-112.2) | 76.7 (63.24-93.02) | Placebo d1 vs. ARVAC Bivalent d1 | ns | >.9999 |
| | | | | | Placebo d1 vs. ARVAC Omicron d1 | ns | >.9999 |
| | | | | | Placebo d1 vs. ARVAC Gamma d1 | ns | >.9999 |
| | | | | | ARVAC Bivalent d1 vs. ARVAC Omicron d1 | ns | >.9999 |
| | | | | | ARVAC Bivalent d1 vs. ARVAC Gamma d1 | ns | >.9999 |
| | | | | | ARVAC Omicron d1 vs. ARVAC Gamma d1 | ns | >.9999 |
| d14 | 77.29 (63.62-93.89) | 962.2 (786.8-1177) | 828.4 (663.6-1034) | 954.2 (769.2-1184) | Placebo d14 vs. ARVAC Bivalent d14 | **** | <.0001 |
| | | | | | Placebo d14 vs. ARVAC Omicron d14 | **** | <.0001 |
| | | | | | Placebo d14 vs. ARVAC Gamma d14 | **** | <.0001 |
| | | | | | ARVAC Bivalent d14 vs. ARVAC Omicron d14 | ns | >.9999 |
| | | | | | ARVAC Bivalent d14 vs. ARVAC Gamma d14 | ns | >.9999 |
| | | | | | ARVAC Omicron d14 vs. ARVAC Gamma d14 | ns | >.9999 |
| **Omicron BA.5** | | | | | | | |
| d1 | 41.82 (34.6-50.55) | 53.82 (43.09-67.21) | 55.57 (45.1-68.47) | 47.82 (39.26-58.25) | Placebo d1 vs. ARVAC Bivalent d1 | ns | >.9999 |
| | | | | | Placebo d1 vs. ARVAC Omicron d1 | ns | .5717 |
| | | | | | Placebo d1 vs. ARVAC Gamma d1 | ns | .6605 |
| | | | | | ARVAC Bivalent d1 vs. ARVAC Omicron d1 | ns | >.9999 |
| | | | | | ARVAC Bivalent d1 vs. ARVAC Gamma d1 | ns | >.9999 |
| | | | | | ARVAC Omicron d1 vs. ARVAC Gamma d1 | ns | >.9999 |
| d14 | 42.93 (35.09-52.53) | 503 (409.8-617.4) | 738.6 (578.4-943.2) | 792.7 (628.4-999.9) | Placebo d14 vs. ARVAC Bivalent d14 | **** | <.0001 |
| | | | | | Placebo d14 vs. ARVAC Omicron d14 | **** | <.0001 |
| | | | | | Placebo d14 vs. ARVAC Gamma d14 | **** | <.0001 |
| | | | | | ARVAC Bivalent d14 vs. ARVAC Omicron d14 | ns | >.9999 |
| | | | | | ARVAC Bivalent d14 vs. ARVAC Gamma d14 | ns | .0898 |

| | | |
|---|---|---|
| ARVAC Omicron d14 vs. ARVAC Gamma d14 | ns | .3704 |

CI, confidence interval; GMT, geometric mean titers; ns, not significant.

[a]Non-parametric two-tailed Kruskal Wallis test for unpaired data and Dunns test for multiple comparisons.

**Table S12.** Comparison of geometric mean titers (GMT) between groups before and after placebo or vaccine administration in Phase III participants >60 years. (N=425).

| SARS-CoV-2 variant and day | Placebo (N=106) GMT (95% CI) | Gamma (N=109) GMT (95% CI) | Omicron (N=108) GMT (95% CI) | Bivalent (N=102) GMT (95% CI) | Comparisons between treatments | Significance | Adjusted *p*-value[a] |
|---|---|---|---|---|---|---|---|
| **Ancestral** | | | | | | | |
| d1 | 171.8 (128, 230.5) | 150.1 (111.9, 201.1) | 208.5 (159.9, 271.8) | 160.2 (121.3, 211.5) | Placebo d1 vs. ARVAC Bivalent d1 | ns | >.9999 |
| | | | | | Placebo d1 vs. ARVAC Omicron d1 | ns | >.9999 |
| | | | | | Placebo d1 vs. ARVAC Gamma d1 | ns | >.9999 |
| | | | | | ARVAC Bivalent d1 vs. ARVAC Omicron d1 | ns | >.9999 |
| | | | | | ARVAC Bivalent d1 vs. ARVAC Gamma d1 | ns | >.9999 |
| | | | | | ARVAC Omicron vs. ARVAC Gamma d1 | ns | .9919 |
| d14 | 192 (142, 259.6) | 1363 (1006, 1847) | 1554 (1205, 2004) | 1837 (1404, 2403) | Placebo d14 vs. ARVAC Bivalent d14 | **** | <.0001 |
| | | | | | Placebo d14 vs. ARVAC Omicron d14 | **** | <.0001 |
| | | | | | Placebo d14 vs. ARVAC Gamma d14 | **** | <.0001 |
| | | | | | ARVAC Bivalent d14 vs. ARVAC Omicron d14 | ns | >.9999 |
| | | | | | ARVAC Bivalent d14 vs. ARVAC Gamma d14 | ns | .8103 |
| | | | | | ARVAC Omicron d14 vs. ARVAC Gamma d14 | ns | >.9999 |
| **Gamma** | | | | | | | |
| d1 | 128.8 (94.95, 174.8) | 105.8 (79.97, 139.9) | 117.8 (90.13, 153.8) | 105.8 (80.32, 139.4) | Placebo d1 vs. ARVAC Bivalent d1 | ns | >.9999 |
| | | | | | Placebo d1 vs. ARVAC Omicron d1 | ns | >.9999 |
| | | | | | Placebo d1 vs. ARVAC Gamma d1 | ns | >.9999 |
| | | | | | ARVAC Bivalent d1 vs. ARVAC Omicron d1 | ns | >.9999 |
| | | | | | ARVAC Bivalent d1 vs. ARVAC Gamma d1 | ns | >.9999 |
| | | | | | ARVAC Omicron vs. ARVAC Gamma d1 | ns | >.9999 |
| d14 | 138.4 (101.8, 188.3) | 819.7 (596.5, 1126) | 855.6 (647.6, 1130) | 1264 (955.9, 1672) | Placebo d14 vs. ARVAC Bivalent d14 | **** | <.0001 |
| | | | | | Placebo d14 vs. ARVAC Omicron d14 | **** | <.0001 |
| | | | | | Placebo d14 vs. ARVAC Gamma d14 | **** | <.0001 |
| | | | | | ARVAC Bivalent d14 vs. ARVAC Omicron d14 | ns | .5724 |
| | | | | | ARVAC Bivalent d14 vs. ARVAC Gamma d14 | ns | .6200 |
| | | | | | ARVAC Omicron d14 vs. ARVAC Gamma d14 | ns | >.9999 |
| **Omicron BA.5** | | | | | | | |
| d1 | 57.64 (42.35, 78.45) | 47.47 (35.13, 64.13) | 64 (48.89, 83.78) | 57.8 (43.6, 76.61) | Placebo d1 vs. ARVAC Bivalent d1 | ns | >.9999 |
| | | | | | Placebo d1 vs. ARVAC Omicron d1 | ns | >.9999 |
| | | | | | Placebo d1 vs. ARVAC Gamma d1 | ns | >.9999 |
| | | | | | ARVAC Bivalent d1 vs. ARVAC Omicron d1 | ns | >.9999 |
| | | | | | ARVAC Bivalent d1 vs. ARVAC Gamma d1 | ns | >.9999 |
| | | | | | ARVAC Omicron vs. ARVAC Gamma d1 | ns | >.9999 |
| d14 | 61.94 (45.57, 84.19) | 336.5 (244, 464) | 728.7 (559.6, 949) | 785.6 (592.4, 1042) | Placebo d14 vs. ARVAC Bivalent d14 | **** | <.0001 |
| | | | | | Placebo d14 vs. ARVAC Omicron d14 | **** | <.0001 |
| | | | | | Placebo d14 vs. ARVAC Gamma d14 | **** | <.0001 |
| | | | | | ARVAC Bivalent d14 vs. ARVAC Omicron d14 | ns | >.9999 |

| | | |
|---|---|---|
| ARVAC Bivalent d14 vs. ARVAC Gamma d14 | ** | .0026 |
| ARVAC Omicron d14 vs. ARVAC Gamma d14 | * | .0105 |

CI, confidence interval; GMT, geometric mean titers; ns, not significant.

[a]Non-parametric two-tailed Kruskal Wallis test for unpaired data and Dunns test for multiple comparisons.

**Table S13.** Comparison of geometric mean fold rises (GMFR) (d1 to d14) of neutralizing antibody titers between groups after placebo or any ARVAC vaccine administration in Phase III participants. N=1053

| SARS-CoV-2 variant | Placebo (N=264) GMFR (95% CI) | Gamma (N=259) GMFR (95% CI) | Omicron (N=265) GMFR (95% CI) | Bivalent (N=265) GMFR (95% CI) | Comparisons between treatments | Significance | Adjusted *p*-value[a] |
|---|---|---|---|---|---|---|---|
| **Ancestral** | | | | | Placebo vs. ARVAC Gamma | **** | <.0001 |
| | | | | | Placebo vs. ARVAC Omicron | **** | <.0001 |
| | 1.0 | 9.6 | 7.5 | 12.1 | Placebo vs. ARVAC Bivalent | **** | <.0001 |
| | (0.9-1.2) | (8.0-11.5) | (6.3-8.9) | (10.2-14.2) | ARVAC Gamma vs. ARVAC Omicron | ns | .4556 |
| | | | | | ARVAC Gamma vs. ARVAC Bivalent | ns | .3344 |
| | | | | | ARVAC Omicron vs. ARVAC Bivalent | ** | .0014 |
| **Gamma** | | | | | Placebo vs. ARVAC Gamma | **** | <.0001 |
| | | | | | Placebo vs. ARVAC Omicron | **** | <.0001 |
| | 1.0 | 10.3 | 8.3 | 12.2 | Placebo vs. ARVAC Bivalent | **** | <.0001 |
| | (0.9-1.1) | (8.5-12.5) | (6.9-10.0) | (10.4-14.4) | ARVAC Gamma vs. ARVAC Omicron | ns | .5031 |
| | | | | | ARVAC Gamma vs. ARVAC Bivalent | ns | .7924 |
| | | | | | ARVAC Omicron vs. ARVAC Bivalent | ** | .0076 |
| **Omicron BA.5** | | | | | Placebo vs. ARVAC Gamma | **** | <.0001 |
| | | | | | Placebo vs. ARVAC Omicron | **** | <.0001 |
| | 1.0 | 8.3 | 12.5 | 15.3 | Placebo vs. ARVAC Bivalent | **** | <.0001 |
| | (0.9-1.2) | (7.0-10.0) | (10.3-15.2) | (12.9-18.2) | ARVAC Gamma vs. ARVAC Omicron | ns | .091 |
| | | | | | ARVAC Gamma vs. ARVAC Bivalent | *** | .0001 |
| | | | | | ARVAC Omicron vs. ARVAC Bivalent | ns | .4176 |

CI, confidence interval; GMFR, geometric mean fold rise.

[a]Non parametric two-tailed Kruskal Wallis test for unpaired data and Dunns test for multiple comparisons.

**Table S14.** Comparison of geometric mean titer ratios (GMTR) between groups after administration of placebo or any ARVAC vaccine in Phase III participants. N=1053

| Comparisons | GMTR | 95% CI | *P*-value |
|---|---|---|---|
| ***All participants*** | | | |
| **Ancestral** | | | |
| Bivalent vs. Gamma | 1.25 | 0.98-1.59 | .067 |
| Bivalent vs. Omicron | 1.32 | 1.04-1.67 | .022 |
| Bivalent vs. Placebo | 11.7 | 9.48-14.50 | <.001 |
| Gamma vs. Omicron | 0.98 | 0.78-1.25 | .899 |
| Gamma vs. Placebo | 8.98 | 7.14-11.30 | <.001 |
| Omicron vs. Placebo | 8.58 | 6.87-10.70 | <.001 |
| **Gamma** | | | |
| Bivalent vs. Gamma | 1.26 | 1.00-1.59 | .048 |
| Bivalent vs. Omicron | 1.34 | 1.06-1.70 | .013 |
| Bivalent vs. Placebo | 12.3 | 10.10-15.10 | <.001 |
| Gamma vs. Omicron | 0.91 | 0.72-1.16 | .454 |
| Gamma vs. Placebo | 9.31 | 7.51-11.60 | <.001 |
| Omicron vs. Placebo | 8.39 | 6.70-10.50 | <.001 |
| **Omicron BA.5** | | | |
| Bivalent vs. Gamma | 1.94 | 1.53-2.46 | <.001 |
| Bivalent vs. Omicron | 1.13 | 0.88-1.44 | .336 |
| Bivalent vs. Placebo | 15.5 | 12.50-19.40 | <.001 |
| Gamma vs. Omicron | 1.71 | 1.34-2.17 | <.001 |
| Gamma vs. Placebo | 7.92 | 6.33-9.91 | <.001 |
| Omicron vs. Placebo | 13.5 | 10.70-17.10 | <.001 |
| ***Participants 18-60 years*** | | | |
| **Ancestral** | | | |
| Bivalent vs. Gamma | 1.16 | 0.86-1.57 | .336 |
| Bivalent vs. Omicron | 1.38 | 1.01-1.87 | .043 |
| Bivalent vs. Placebo | 12.4 | 9.59-16.10 | <.001 |
| Gamma vs. Omicron | 0.91 | 0.67-1.24 | .554 |
| Gamma vs. Placebo | 10.7 | 8.12-14.0 | <.001 |
| Omicron vs. Placebo | 9.04 | 6.85-11.9 | <.001 |
| **Gamma** | | | |
| Bivalent vs. Gamma | 1.04 | 0.79-1.38 | .773 |
| Bivalent vs. Omicron | 1.26 | 0.95-1.68 | .107 |
| Bivalent vs. Placebo | 12.5 | 9.86-15.8 | <.001 |
| Gamma vs. Omicron | 0.85 | 0.64-1.13 | .271 |
| Gamma vs. Placebo | 11.8 | 9.12-15.2 | <.001 |
| Omicron vs. Placebo | 9.65 | 7.45-12.50 | <.001 |
| **Omicron BA.5** | | | |
| Bivalent vs. Gamma | 1.71 | 1.28-2.28 | <.001 |
| Bivalent vs. Omicron | 1.14 | 0.82-1.58 | .449 |
| Bivalent vs. Placebo | 16.6 | 12.70-21.9 | <.001 |
| Gamma vs. Omicron | 1.46 | 1.07-1.99 | .016 |
| Gamma vs. Placebo | 9.9 | 7.62-12.9 | <.001 |
| Omicron vs. Placebo | 15.4 | 11.30-21.0 | <.001 |
| ***Participants >60 years*** | | | |
| **Ancestral** | | | |
| Bivalent vs. Gamma | 1.4 | 0.95-2.06 | .089 |
| Bivalent vs. Omicron | 1.28 | 0.90-1.82 | .173 |
| Bivalent vs. Placebo | 10.6 | 7.36-15.10 | <.001 |
| Gamma vs. Omicron | 1.09 | 0.74-1.58 | .668 |
| Gamma vs. Placebo | 7.29 | 4.94-10.7 | <.001 |
| Omicron vs. Placebo | 7.72 | 5.39-11.1 | <.001 |
| **Gamma** | | | |
| Bivalent vs. Gamma | 1.67 | 1.13-2.49 | .011 |

| | | | |
|---|---|---|---|
| Bivalent vs. Omicron | 1.56 | 1.06-2.29 | .023 |
| Bivalent vs. Placebo | 11.5 | 8.13-16.4 | <.001 |
| Gamma vs. Omicron | 1.01 | 0.67-1.52 | .961 |
| Gamma vs. Placebo | 6.68 | 4.59-9.72 | <.001 |
| Omicron vs. Placebo | 6.22 | 4.25-9.13 | <.001 |
| **Omicron BA.5** | | | |
| Bivalent vs. Gamma | 2.38 | 1.59-3.58 | <.001 |
| Bivalent vs. Omicron | 1.12 | 0.77-1.62 | .553 |
| Bivalent vs. Placebo | 13.3 | 9.18-19.2 | <.001 |
| Gamma vs. Omicron | 2.13 | 1.44-3.14 | <.001 |
| Gamma vs. Placebo | 5.45 | 3.67-8.10 | <.001 |
| Omicron vs. Placebo | 11.5 | 8.02-16.4 | <.001 |

CI, confidence interval; GMTR, geometric mean titers ratio.

**Table S15.** Local adverse events according to vaccine variant in Phase II and Phase III participants.

| | | Bivalent (n=581) | | | | Gamma (n=800) | | | | Omicron (n=580) | | | | Total vaccine | *P*-value[b] |
|---|---|---|---|---|---|---|---|---|---|---|---|---|---|---|---|
| | | Grade 1 | Grade 2 | Grade 3 | Total[a] | Grade 1 | Grade 2 | Grade 3 | Total[a] | Grade 1 | Grade 2 | Grade 3 | Total[a] | n=1961 | |
| **Pain** | n | 262 | 10 | | 272 | 304 | 11 | | 315 | 244 | 10 | | 254 | 841 | .019 |
| | %[c] | 96.3 | 4.0 | | 46.8 | 96.5 | 3.5 | | 39.4 | 96.1 | 3.9 | | 43.8 | 42.9 | |
| **Sensitivity / discomfort** | n | 181 | 25 | 1 | 207 | 217 | 29 | 2 | 248 | 167 | 15 | 1 | 183 | 638 | .161 |
| | %[c] | 87.4 | 12.1 | 0.5 | 35.6 | 87.5 | 11.7 | 0.8 | 31.0 | 91.3 | 8.2 | 0.5 | 31.6 | 32.5 | |
| **Erythema / redness** | n | 20 | 1 | | 21 | 25 | | | 25 | 29 | 2 | | 31 | 77 | .1 |
| | %[c] | 95.2 | 4.8 | | 3.6 | 100.0 | | | 3.1 | 93.5 | 6.5 | | 5.3 | 3.9 | |
| **Swelling / induration** | n | 49 | | | 49 | 55 | 1 | | 56 | 51 | 2 | | 53 | 158 | .327 |
| | %[c] | 100.0 | | | 8.4 | 98.2 | 1.8 | | 7.0 | 96.2 | 3.8 | | 9.1 | 8.1 | |
| **Itching** | n | 18 | | | 18 | 21 | | | 21 | 15 | | | 15 | 54 | .832 |
| | %[c] | 100.0 | | | 3.1 | 100.0 | | | 2.6 | 100.0 | | | 2.6 | 2.8 | |

[a] Percentage according to the total number of group participants.
[b] Chi-square test.
[c] Intensity grade percentage according to the total number of events.

**Table S16.** Systemic adverse events according to vaccine variant in Phase II and Phase III participants.

| | | Bivalent (n=581) | | | | | Gamma (n=800) | | | | Omicron (n=580) | | | | P-value[b] |
|---|---|---|---|---|---|---|---|---|---|---|---|---|---|---|---|
| | | Grade 1 | Grade 2 | Grade 3 | Grade 4 | Total[a] | Grade 1 | Grade 2 | Grade 3 | Total[a] | Grade 1 | Grade 2 | Grade 3 | Total[a] | |
| **Diarrhea** | n | 15 | 1 | 1 | | 17 | 13 | 1 | | 14 | 15 | 1 | | 16 | .294 |
| | %[c] | 88.2 | 5.9 | 5.9 | | 2.9 | 92.9 | 7.1 | | 1.8 | 93.8 | 6.3 | | 2.8 | |
| **Headache** | n | 62 | 13 | 2 | | 77 | 68 | 17 | | 85 | 73 | 7 | | 80 | .153 |
| | %[c] | 80.5 | 16.9 | 2.6 | | 13.3 | 80.0 | 20.0 | | 10.6 | 91.3 | 8.8 | | 13.8 | |
| **Joint pain** | n | 13 | | 1 | | 14 | 21 | 5 | | 26 | 14 | 2 | | 16 | .643 |
| | %[c] | 92.9 | | 7.1 | | 2.4 | 80.8 | 19.2 | | 3.3 | 87.5 | 12.5 | | 2.8 | |
| **Muscle pain/ myalgia** | n | 31 | 3 | 1 | | 35 | 38 | 7 | | 45 | 34 | 5 | | 39 | .699 |
| | %[c] | 88.6 | 8.6 | 2.9 | | 6.0 | 84.4 | 15.6 | | 5.6 | 87.2 | 12.8 | | 6.7 | |
| **Chills** | n | 13 | 3 | 1 | | 17 | 10 | 2 | | 12 | 11 | 2 | | 13 | .192 |
| | %[c] | 76.5 | 17.6 | 5.9 | | 2.9 | 83.3 | 16.7 | | 1.5 | 84.6 | 15.4 | | 2.2 | |
| **Fatigue/tiredness/ decay** | n | 53 | 9 | 2 | 1 | 65 | 90 | 9 | | 99 | 63 | 5 | | 68 | .793 |
| | %[c] | 81.5 | 13.8 | 3.1 | 1.5 | 11.2 | 90.9 | 9.1 | | 12.4 | 92.6 | 38.5 | | 11.7 | |
| **Fever** | n | 6 | 1 | 1 | | 8 | 9 | 5 | | 14 | 7 | | | 7 | .691 |
| | %[c] | 75.0 | 12.5 | 12.5 | | 1.4 | 64.3 | 35.7 | | 1.8 | 100.0 | | | 1.2 | |
| **Nausea** | n | 11 | 1 | | | 12 | 8 | 1 | | 9 | 13 | | | 13 | .224 |
| | %[c] | 91.7 | 8.3 | | | 2.1 | 88.9 | 11.1 | | 1.1 | 100.0 | | | 2.2 | |
| **Palpitations** | n | 6 | 1 | | | 7 | 8 | 1 | | 9 | 1 | 1 | | 2 | .223 |
| | %[c] | 85.7 | 14.3 | | | 1.2 | 88.9 | 11.1 | | 1.1 | 50.0 | 50.0 | | 0.3 | |
| **Drowsiness** | n | 49 | 13 | 2 | | 64 | 79 | 8 | | 87 | 68 | 3 | | 71 | .704 |
| | %[c] | 76.6 | 20.3 | 3.1 | | 11.0 | 90.8 | 9.2 | | 10.9 | 95.8 | 4.2 | | 12. | |
| **Vomiting** | n | 2 | | | | 2 | 3 | | | 3 | 1 | | | 1 | .782 |
| | %[c] | 100.0 | | | | 0.3 | 100.0 | | | 0.4 | 100.0 | | | 0.2 | |

[a] Percentage according to the total number of group participants.
[b] Chi-square test.
[c] Intensity grade percentage according to the total number of events.

#### Supplementary Figures

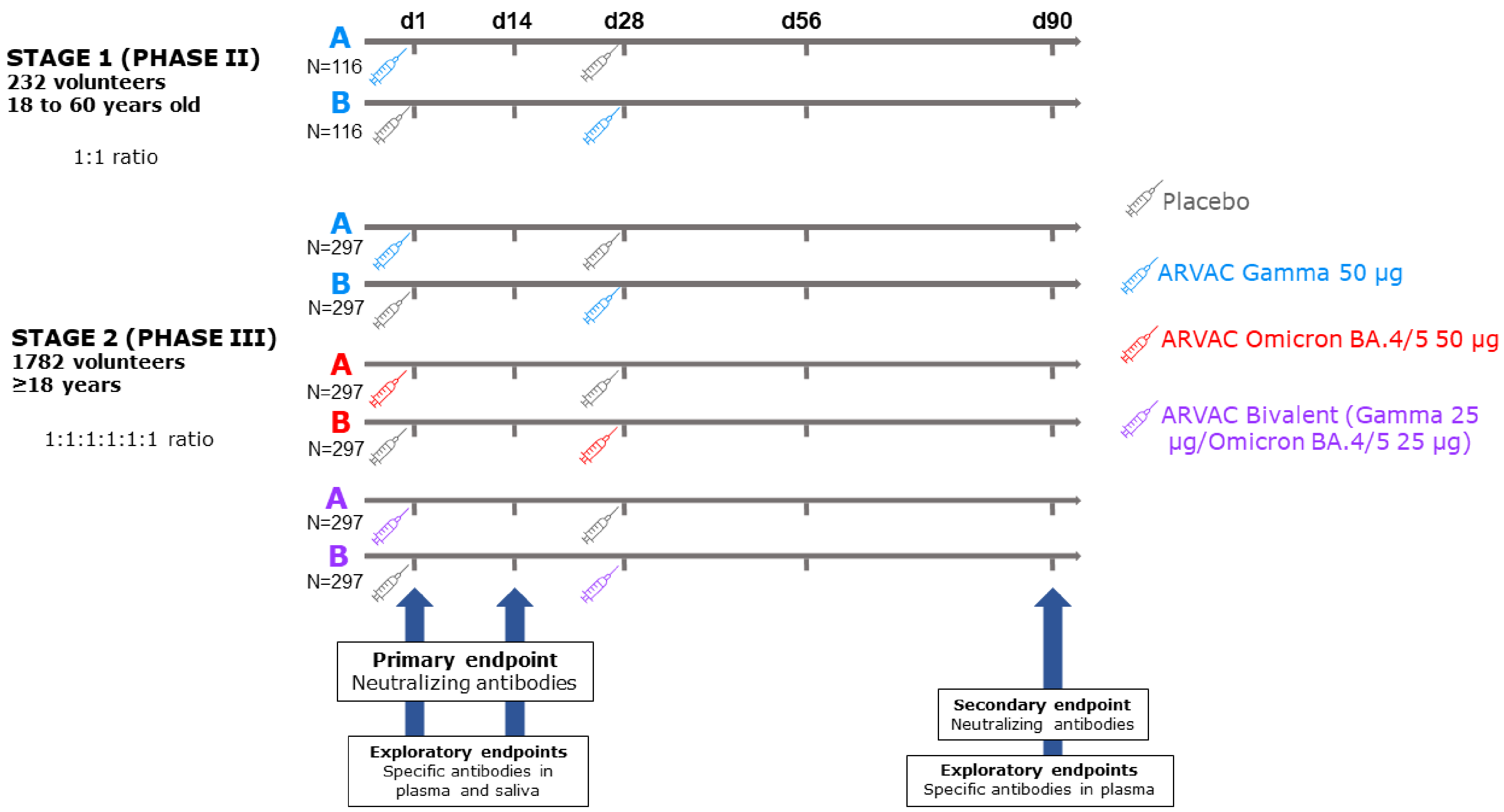


**Figure S1.** Study design.

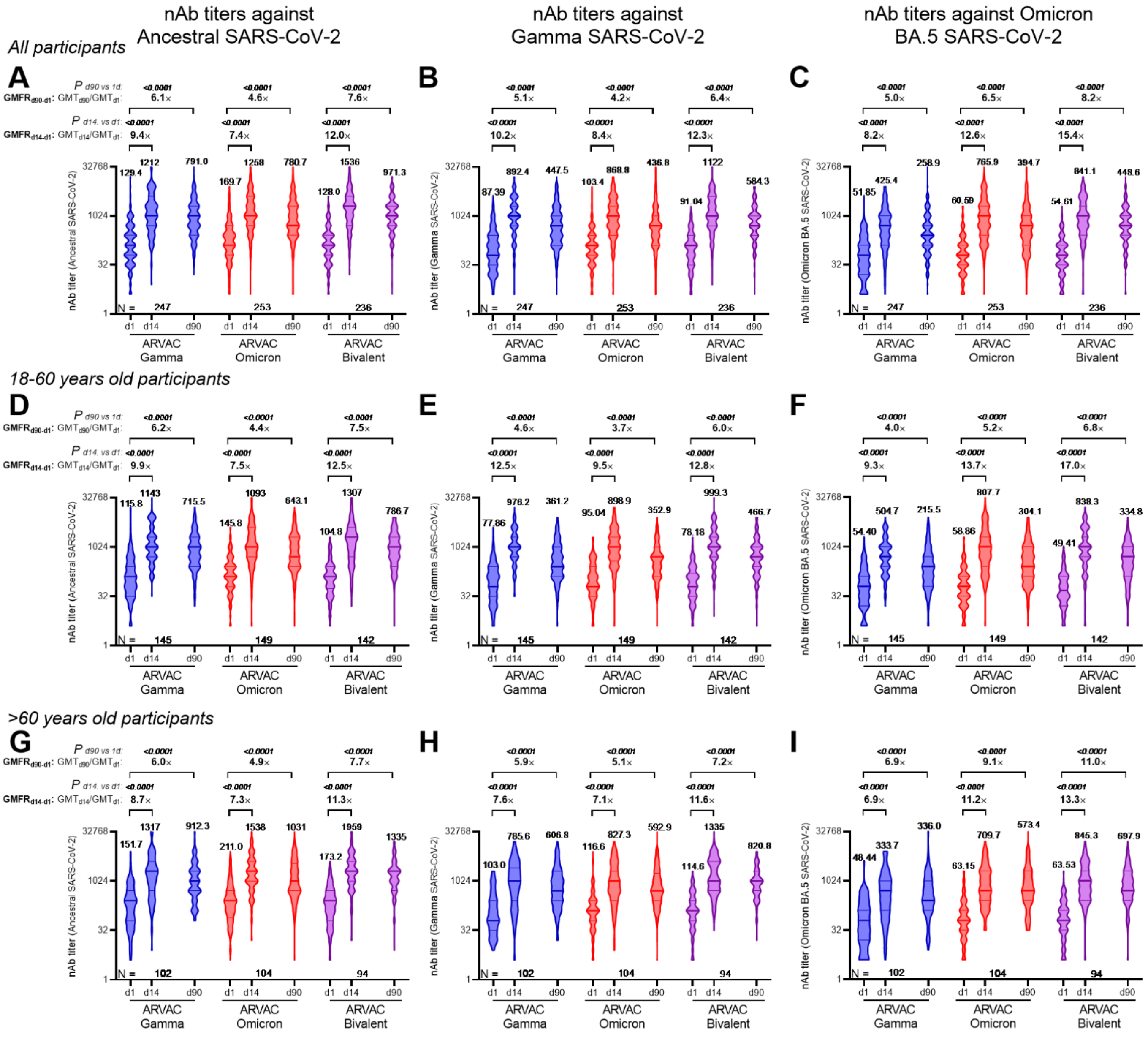

**Figure S2.** Titers of neutralizing antibodies to SARS-CoV-2 (A, D, G) Ancestral, (B, E, H) Gamma, and (C, F, I) Omicron BA.5 variants before (day 1, d1) and at days 14 (d14) and 90 (d90) after administration of ARVAC Gamma (n=247), Omicron B4.5 (n=253), and bivalent (n=236) vaccine versions in (A-C) all participants, (D-F) participants 18-60 years, and (G-I) participants >60 years. The thick horizontal lines in the violin plots represent geometric mean titers (GMT) and the value is indicated above the plots. Geometric mean fold rises (GMFR) after (d14 and d90) administration and *p*-values comparing neutralizing antibody levels with respect to d1 are indicated. *P*-values were calculated using the non-parametric two-tailed paired Friedman test and the Dunns test. GMFR, geometric mean fold rise; GMT, geometric mean titers; nAbs, neutralizing antibody titers.

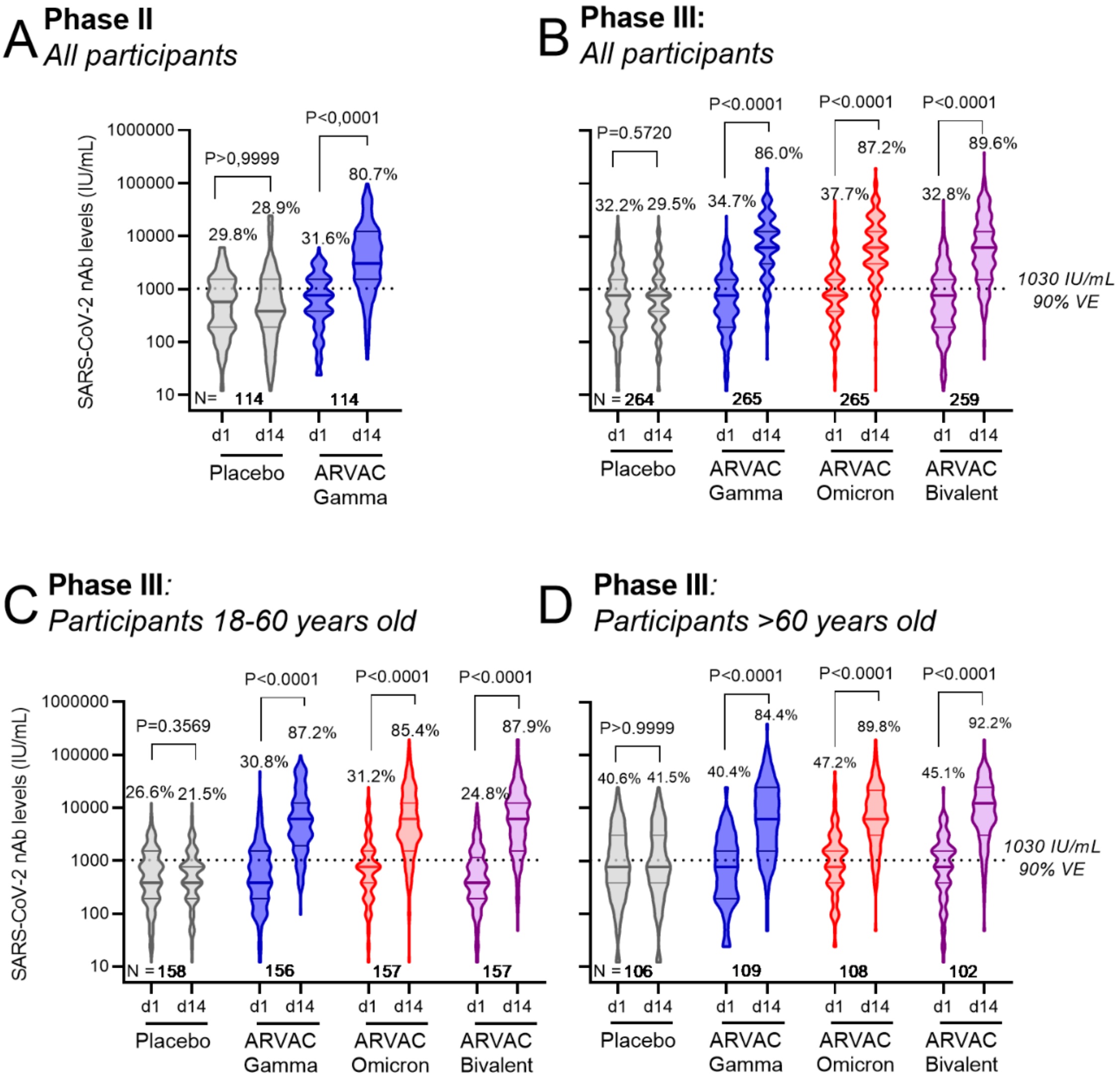


**Figure S3.** Levels of neutralizing antibodies to SARS-CoV-2 Ancestral variant expressed as normalized units (UI/mL) before (day 1) and after (day 14) vaccine and placebo administration. The graphs show data from participants in Phase II (A) and Phase III, including all participants (B), participants 18-60 years (C), and participants >60 years (D) receiving the ARVAC Gamma, ARVAC Omicron, and ARVAC bivalent vaccine versions. The thick horizontal line in the violin plots represent the geometric mean. The horizontal dotted line represents the proposed threshold of 1030 UI/mL for a 90% protection against symptomatic infection and the percentage of participants above the threshold are shown above each plot. Differences between proportions before and after administration were calculated using the Chi-square test. Ns, not significant: $p > 0.05$; ****, $p < 0.0001$. nAb, neutralizing antibodies.

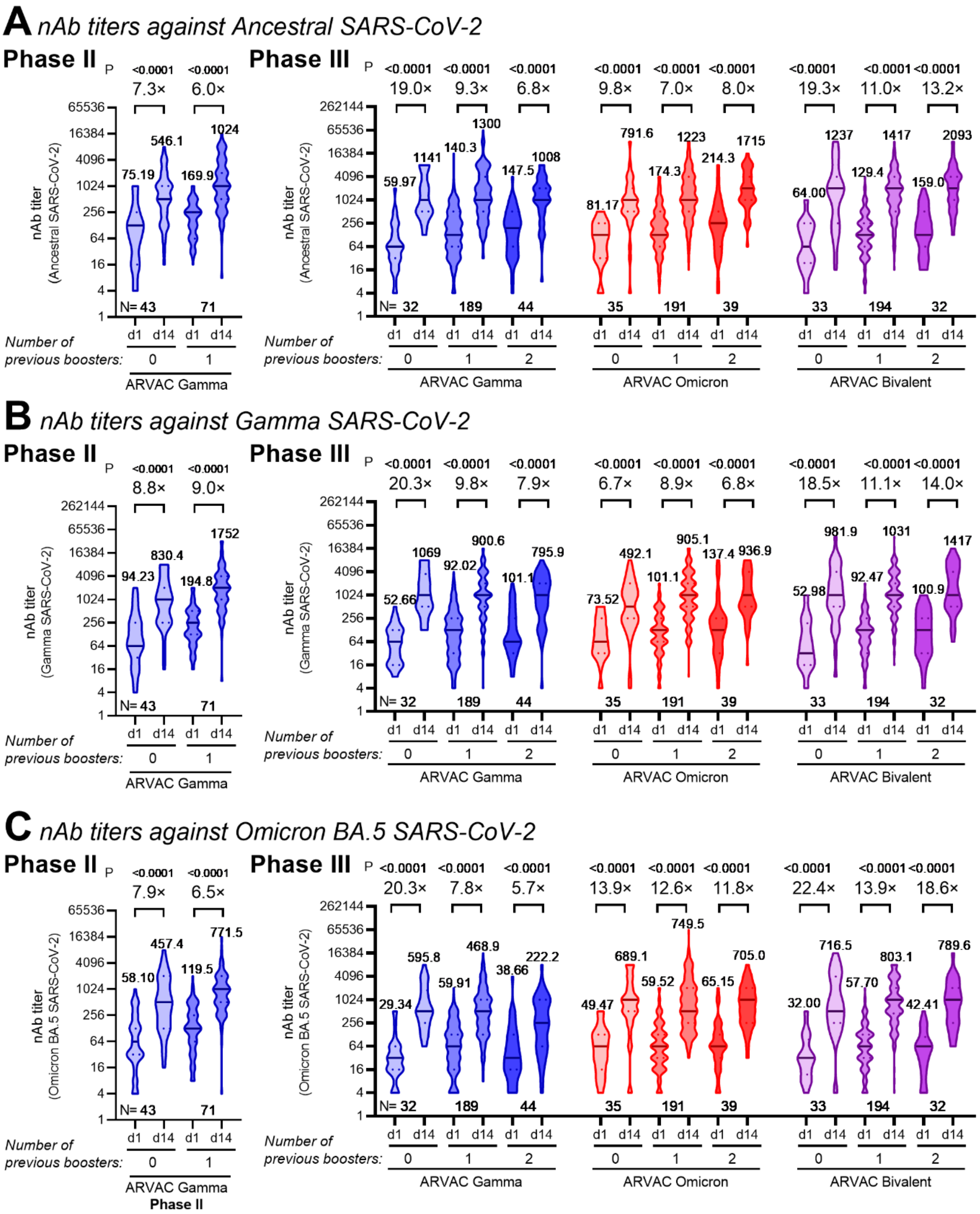


**Figure S4.** Titers of neutralizing antibodies to SARS-CoV-2 variants before (day 1) and after (day 14) vaccine and placebo administration according to previous booster administration in Phase II and Phase III participants. The graphs represent the titers to Ancestral (A), Gamma (B), and Omicron BA.5 (C) variants. The thick horizontal line in the violin plots represent the geometric mean, which is indicated above the plots. Geometric mean fold rises (GMFR) and $p$-values before and after administration are also indicated. $P$-values were calculated using the non-parametric paired Wilcoxon test. nAb, neutralizing antibody.

#### ARVAC Gamma: Phase II

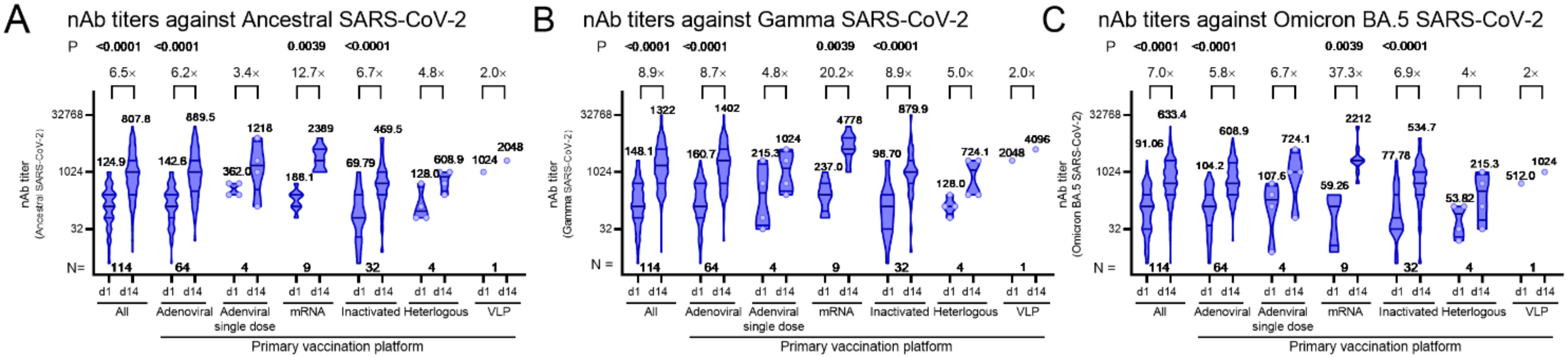


**Figure S5.** Titers of neutralizing antibodies to SARS-CoV-2 Ancestral (A), Gamma (B), and Omicron BA.5 (C) variants in Phase II participants before (day 1) and after (day 14) vaccine and placebo administration according to previous primary vaccination schemes. The vaccination schemes considered were adenovirus, adenovirus single dose, mRNA, inactivated virus, heterologous vaccination, and virus like particle. The thick horizontal lines in the violin plots represent the geometric mean, which is indicated above the plots. Geometric mean fold rises (GMFR) and *p*-values before and after administration are also indicated. *P*-values were calculated using the non-parametric paired Wilcoxon test. nAb, neutralizing antibody; VLP, virus-like particle.

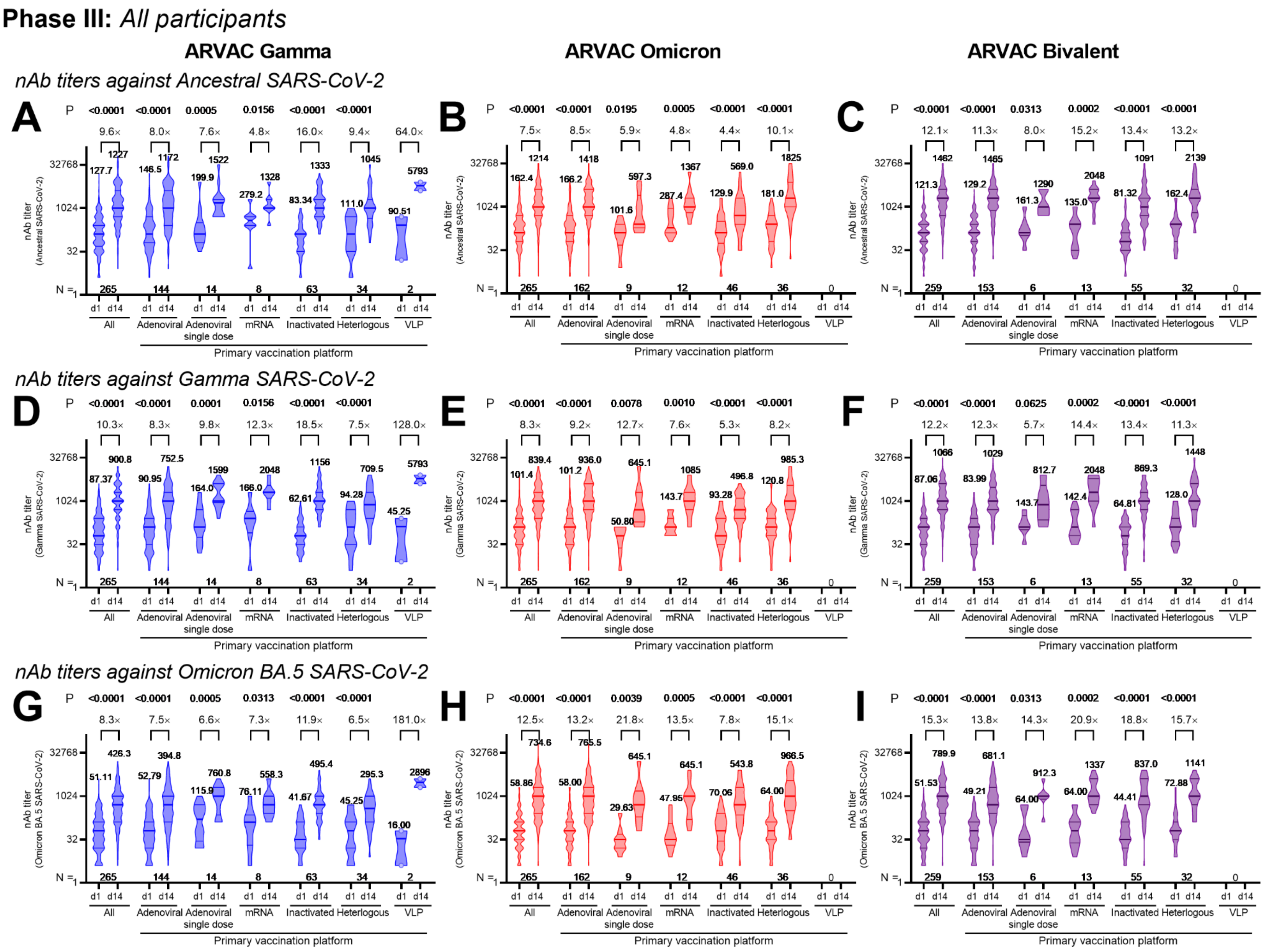
Phase III: All participants
ARVAC Gamma
ARVAC Omicron
ARVAC Bivalent
nAb titers against Ancestral SARS-CoV-2
nAb titers against Gamma SARS-CoV-2
nAb titers against Omicron BA.5 SARS-CoV-2
A
B
C
D
E
F
G
H
I
P
nAb titer (Ancestral SARS-CoV-2)
nAb titer (Gamma SARS-CoV-2)
nAb titer (Omicron BA.5 SARS-CoV-2)
N =
d1 d14
All
Adenoviral
Adenoviral single dose
mRNA
Inactivated
Heterlogous
VLP
Primary vaccination platform

**Figure S6**. Titers of neutralizing antibodies to SARS-CoV-2 Ancestral (A-C), Gamma (D-F), and Omicron BA.5 (G-I) variants in all Phase III participants before (day 1) and after (day 14) vaccine administration according to previous primary vaccination schemes. The graphs represent the titers after ARVAC Gamma (A,D,G), Omicron BA.4/5 (B,E,H), and bivalent (C,F,I) vaccines administration. The vaccination schemes considered were adenovirus, adenovirus single dose, mRNA, inactivated virus, heterologous vaccination, and virus like particle. The thick horizontal lines in the violin plots represent the geometric mean, which is indicated above the plots. Geometric mean fold rises (GMFR) and *p*-values before and after administration are also indicated. *P*-values were calculated using the non-parametric paired Wilcoxon test. nAb, neutralizing antibodies; VLP, virus-like particle.

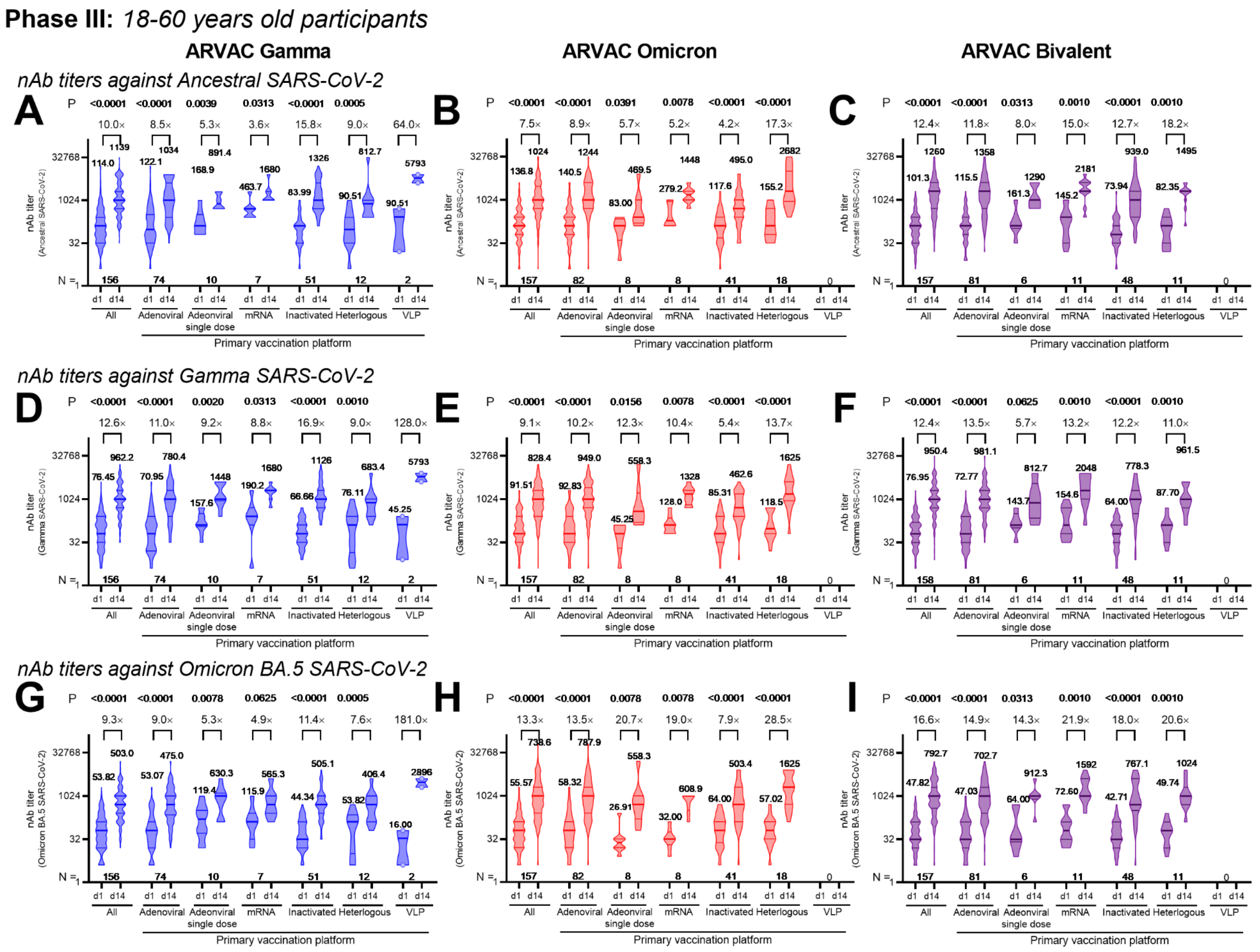

Phase III: 18-60 years old participants
ARVAC Gamma
ARVAC Omicron
ARVAC Bivalent
nAb titers against Ancestral SARS-CoV-2
nAb titers against Gamma SARS-CoV-2
nAb titers against Omicron BA.5 SARS-CoV-2
Primary vaccination platform

**Figure S7**. Titers of neutralizing antibodies to SARS-CoV-2 Ancestral (A-C), Gamma (D-F), and Omicron BA.5 (G-I) variants in 18-60 years Phase III participants before (day 1) and after (day 14) vaccine administration according to previous primary vaccination schemes. The graphs represent the titers after ARVAC Gamma (A,D,G), Omicron BA.4/5 (B,E,H), and bivalent (C,F,I) vaccines administration. The vaccination schemes considered were adenovirus, adenovirus single dose, mRNA, inactivated virus, heterologous vaccination, and virus like particle. The thick horizontal lines in the violin plots represent the geometric mean, which is indicated above the plots. Geometric mean fold rises (GMFR) and *p*-values before and after administration are also indicated. *P*-values were calculated using the non-parametric paired Wilcoxon test. nAb, neutralizing antibodies. VLP, virus-like particle.

**Phase III:** *Participants >60*

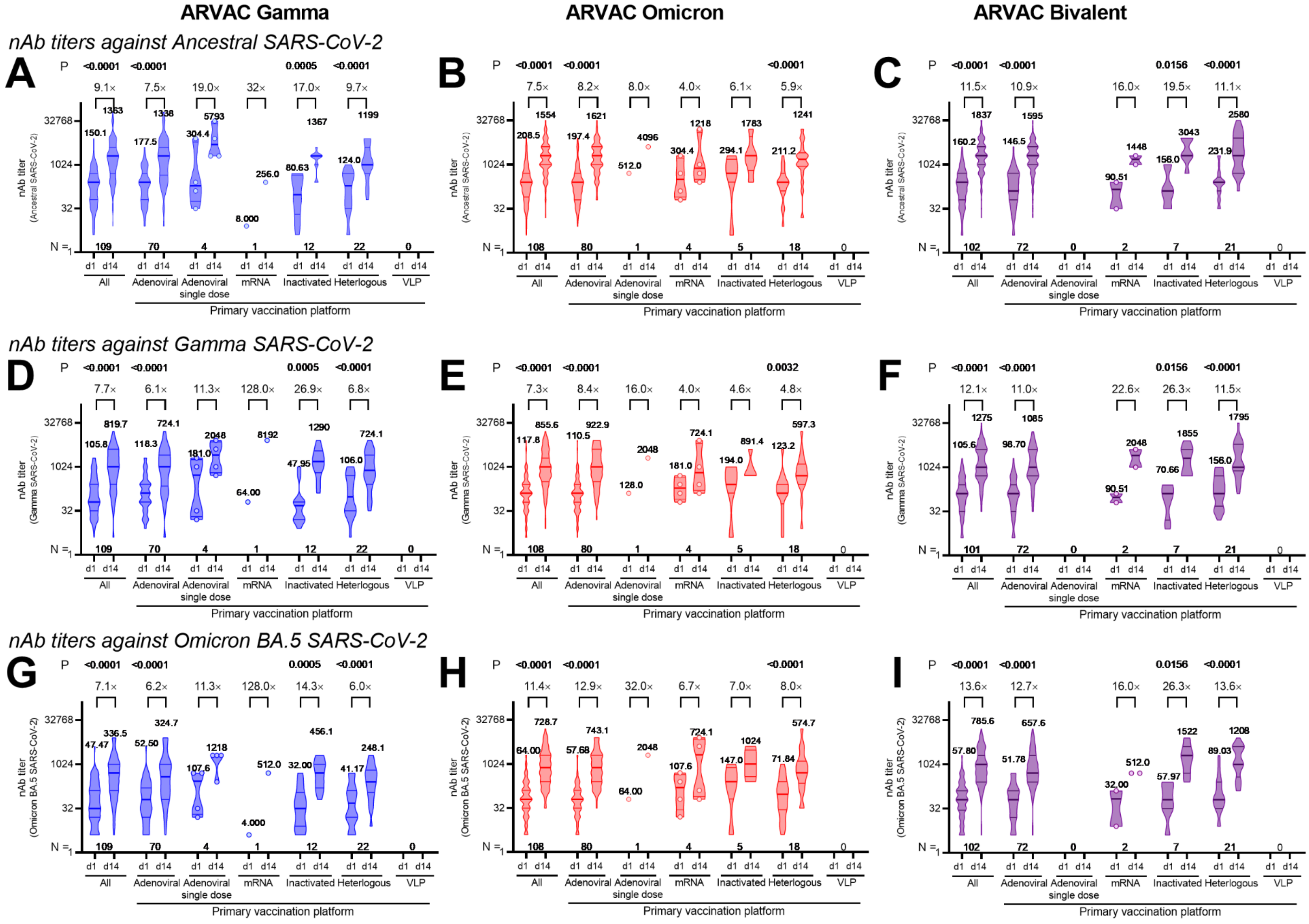

**Figure S8**. Titers of neutralizing antibodies to SARS-CoV-2 Ancestral (A-C), Gamma (D-F), and Omicron BA.5 (G-I) variants in Phase III participants >60 years before (day 1) and after (day 14) vaccine administration according to previous primary vaccination schemes. The graphs represent the titers after ARVAC Gamma (A,D,G), Omicron BA.4/5 (B,E,H), and bivalent (C,F,I) vaccines administration. The vaccination schemes considered were adenovirus, adenovirus single dose, mRNA, inactivated virus, heterologous vaccination, and virus like particle. The thick horizontal lines in the violin plots represent the geometric mean, which is indicated above the plots. Geometric mean fold rises (GMFR) and *p*-values before and after administration are also indicated. *P*-values were calculated using the non-parametric paired Wilcoxon test. nAb, neutralizing antibodies; VLP, virus-like particle.

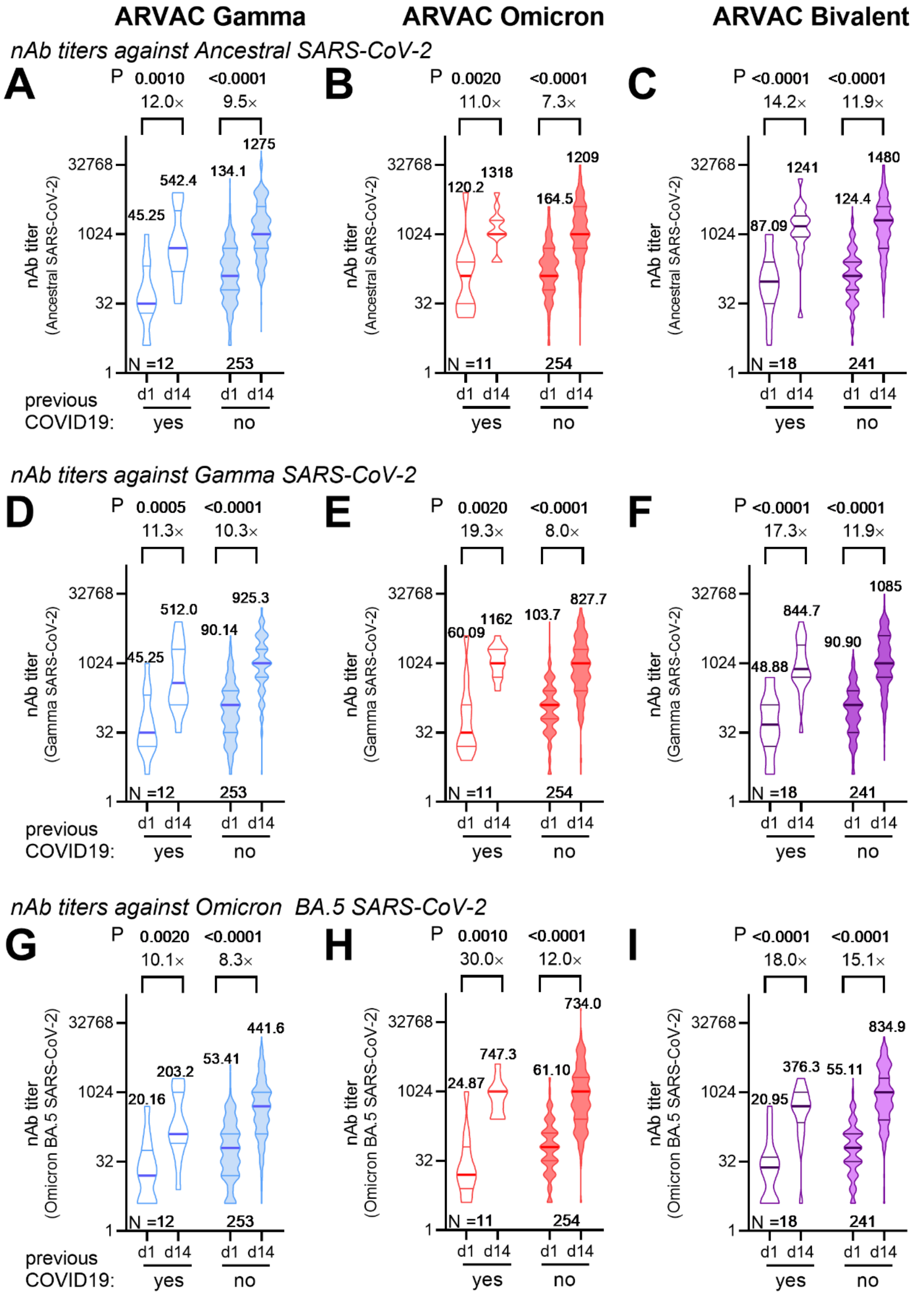


**Figure S9.** Neutralizing antibody titers to SARS-CoV-2 Ancestral (A-C), Gamma (D-F), and Omicron BA.5 (G-I) variants in Phase III participants before (day 1) and after (day 14) vaccine administration according to previous history of COVID-19 before study inclusion based on diagnostic or on seropositivity to SARS-CoV-2 nucleoprotein. The graphs represent the titers after ARVAC Gamma (A,D,G), Omicron BA.4/5 (B,E,H), and

Bivalent (C,F,I) vaccines administration. The thick horizontal lines in the violin plots represent the geometric mean, which is indicated above the plots. Geometric mean fold rises (GMFR) and *p*-values before and after administration are also indicated. *P*-values were calculated using the non-parametric paired Wilcoxon test. nAb, neutralizing antibodies.

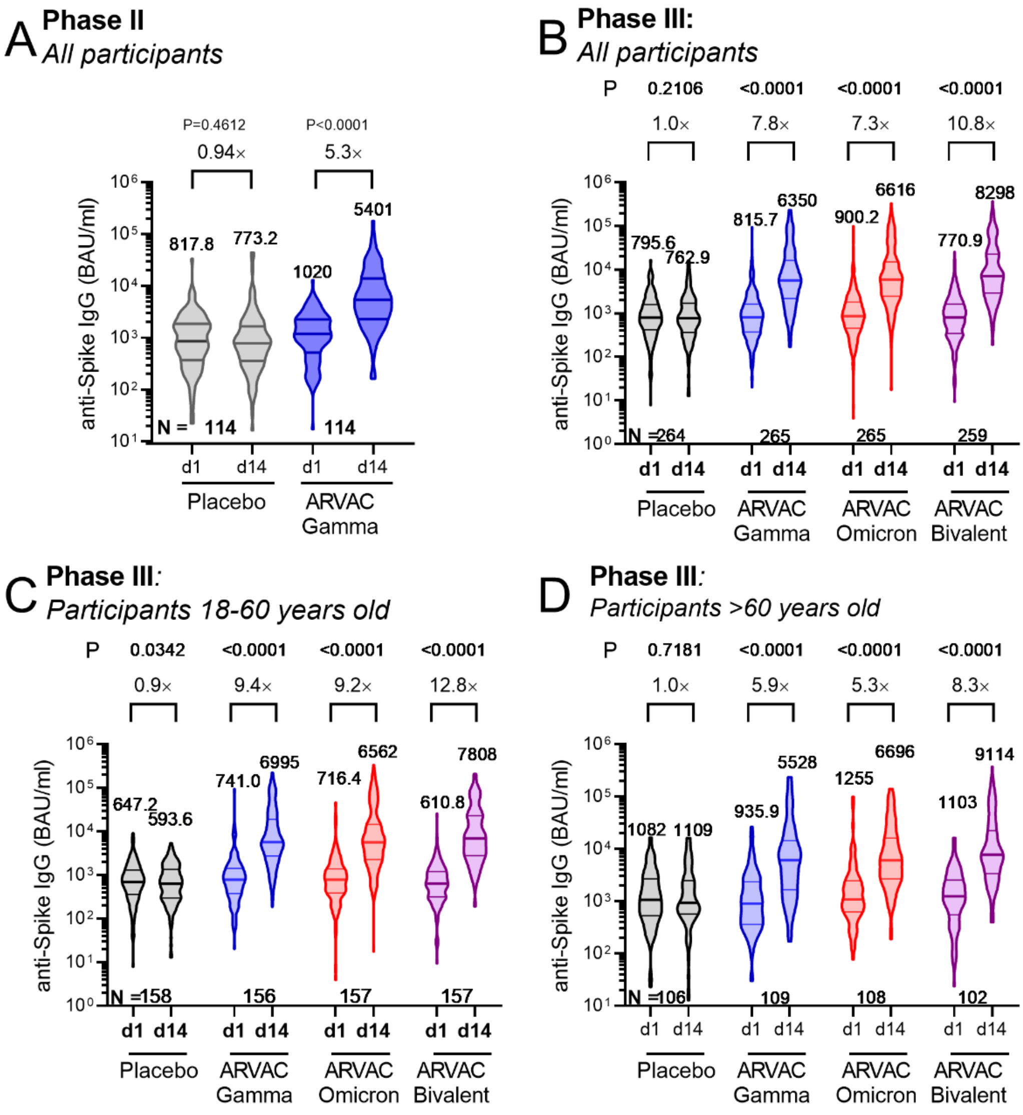


**Figure S10.** Titers of anti-spike-specific IgG antibodies in Phase II (A) and Phase III participants (B-D) before (day 1) and after (day 14) vaccine and placebo administration. The graphs represent data from all participants (B), participants 18-60 years (C), and participants >60 years (D). Each dot represents one data point and the thick horizontal lines in the violin plots represent the geometric mean, which is indicated above the plots. Geometric mean fold rises (GMFR) and *p*-values before and after administration are also indicated. *P*-values were calculated using the non-parametric paired Wilcoxon test.

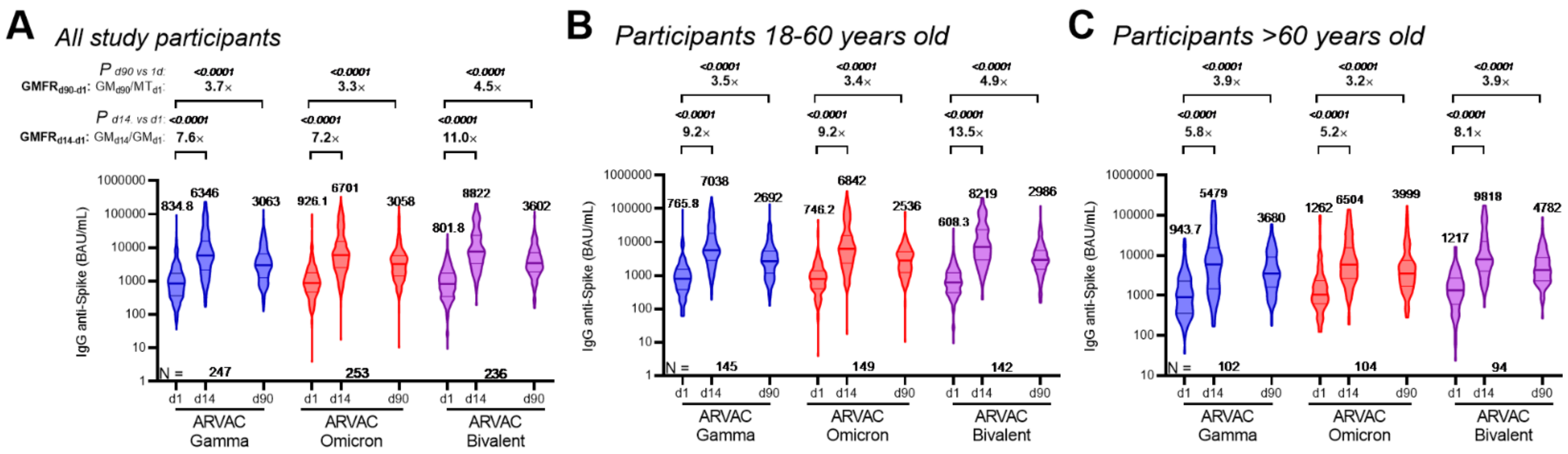


**Figure S11.** Levels of anti-spike-specific IgG antibodies before (day 1, d1) and at days 14 (d14) and 90 (d90) after ARVAC Gamma, Omicron BA.5, and bivalent vaccine administration. The violin plots represent overall data from Phase III participants (A) and according to age group: 18-60 years (B) and >60 years (C). The thick horizontal lines in the violin plots represent the geometric mean, which is indicated above the plots. Geometric mean fold rises (GMFR) and *p*-values before and after administration are also indicated. *P*-values were calculated using the non-parametric paired Wilcoxon test.

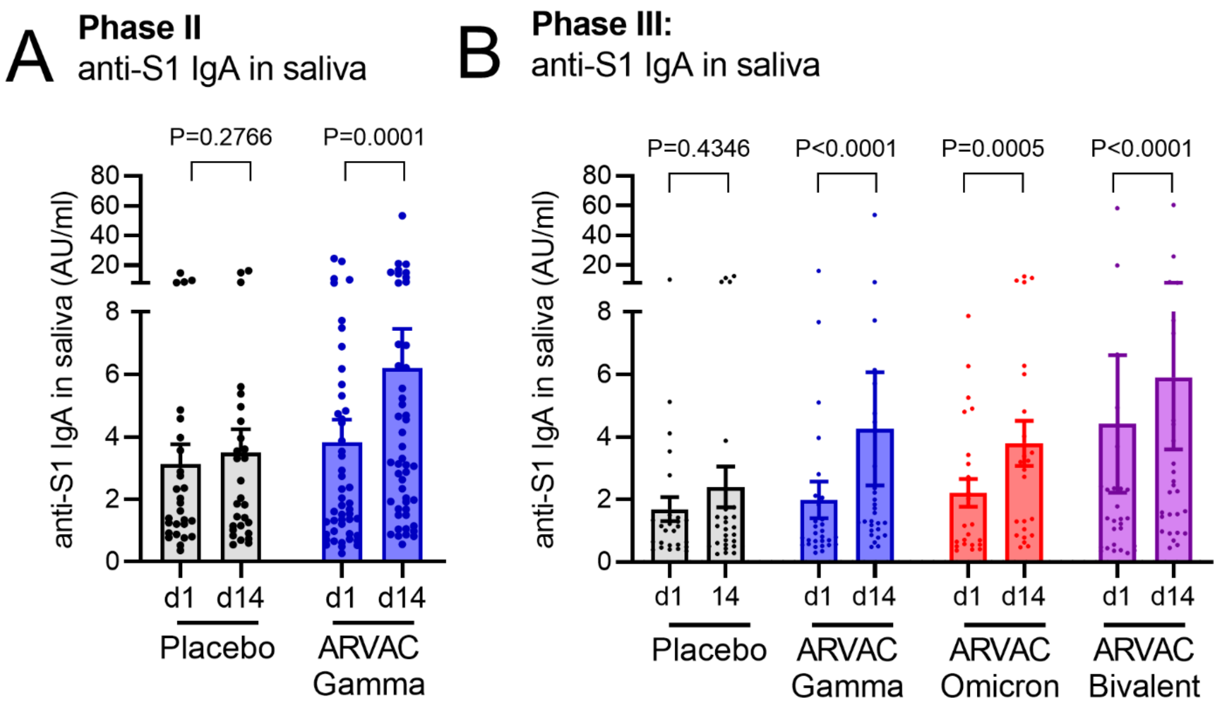


**Figure S12.** Titers of anti-spike-specific IgA antibodies in saliva before (day 1) and after (day 14) vaccine and placebo administration, expressed in arbitrary units. The column graphs represent the mean values from Phase II (A) and Phase III participants (B). Error bars represent the standard deviation. The *p*-values comparing values before and after administration were calculated using the non-parametric paired Wilcoxon test. ***, $p < 0.001$; ****, $p < 0.0001$; ns, not significant ($p > 0.05$).

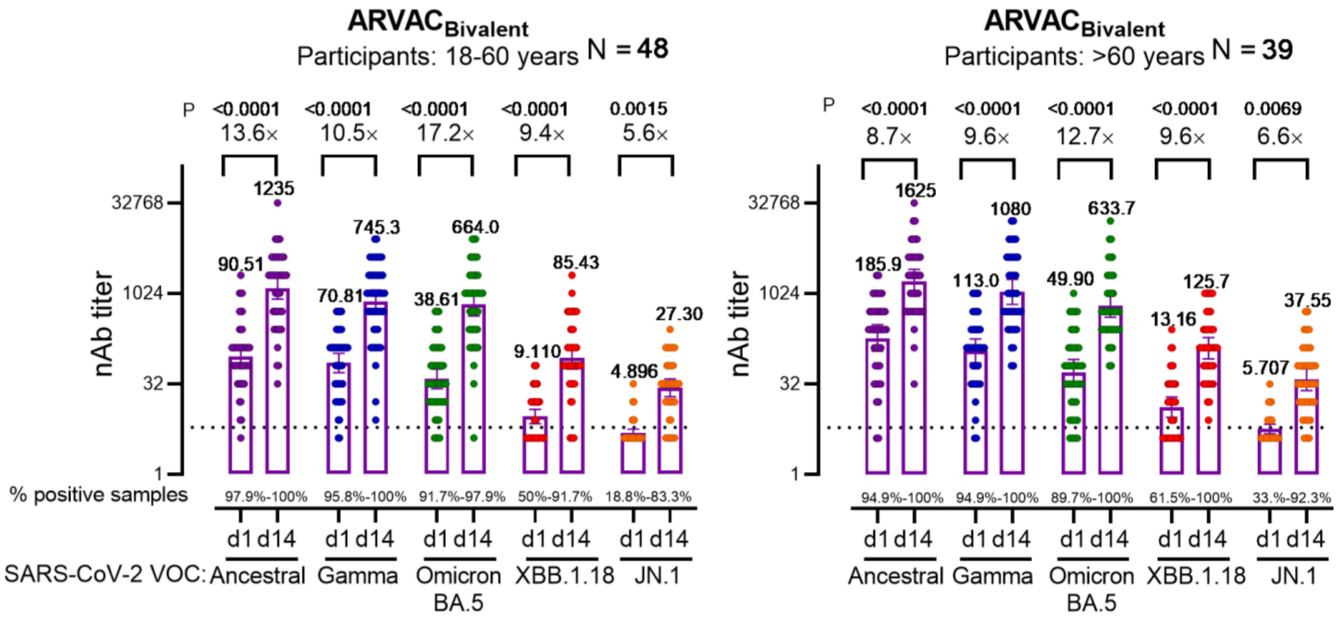


**Figure S13.** Titers of neutralizing antibodies to SARS-CoV-2 Ancestral, Gamma and Omicron subvariants: BA.5, XBB.1.18 and JN.1 before (day 1, d1) and at days 14 (d14) after administration of ARVAC$_{\mathrm{Bivalent}}$ in study participant aged 18-60 (n=48) or study participants >60 years old (n=39). The geometric mean titer (GMT) value is indicated above the bars. Geometric mean fold rises (GMFR) from before (d1) to after (d14) administration and *p*-values comparing levels with respect to d1 are indicated. *P*-values were calculated using the non-parametric two-tailed paired Friedman test followed by the Dunns test. The percentage of positive samples (samples with nAb titer >5) are depicted for each dataset. nAb, neutralizing antibodies.
